# Medication and pharmacogenomic effects on cross-sectional symptom severity and cognitive ability in schizophrenia

**DOI:** 10.1101/2024.11.20.24317526

**Authors:** Siobhan K. Lock, Djenifer B. Kappel, Michael J. Owen, James T.R. Walters, Michael C. O’Donovan, Antonio F. Pardiñas, Sophie E. Legge

**Author notes:** Antonio F. Pardiñas //.

## Abstract

**Background:** People with schizophrenia differ in the type and severity of symptoms experienced, as well as their response to medication. A better understanding of the factors that influence this heterogeneity is necessary for the development of individualised patient care. Here, we sought to investigate the relationships between phenotypic severity and both medication and pharmacogenomic variables in a cross-sectional sample of people with schizophrenia or schizoaffective disorder depressed type.

**Methods:** Confirmatory factor analysis derived five dimensions relating to current symptoms (positive symptoms, negative symptoms of diminished expressivity, negative symptoms of reduced motivation and pleasure, depression and suicide) and cognitive ability in participants prescribed with antipsychotic medication. Linear models were fit to test for associations between medication and pharmacogenomic variables with dimension scores in the full sample (N = 585), and in a sub-sample of participants prescribed clozapine (N = 215).

**Outcomes:** Lower cognitive ability was associated with higher chlorpromazine-equivalent daily antipsychotic dose and with the prescription of clozapine and anticholinergic medication. We also found associations between pharmacogenomics-inferred cytochrome P450 (CYP) enzyme activity and symptom dimensions. Increased genotype-predicted CYP2C19 and CYP3A5 activity were associated with reduced severity of positive and negative symptoms, respectively. Faster predicted CYP1A2 activity was associated with higher cognitive dimension scores in people taking clozapine.

**Interpretation:** Our results confirm the importance of taking account of medication history (and particularly antipsychotic type and dose) in assessing potential causes of cognitive impairment or poor functioning in patients with schizophrenia. We also highlight the potential for pharmacogenomic variation to be a useful tool to help guide drug prescription, although these findings require further validation.

## Introduction

Schizophrenia is characterised by positive, negative, and disorganised symptoms alongside affective features and cognitive deficits. The presentation of schizophrenia is heterogeneous in the range and severity of symptoms and in the degree to which those respond to antipsychotic treatment.

Heterogeneity in treatment response has been associated with pre-morbid functioning, initial symptom severity, age of onset, duration of untreated psychosis, as well as comorbid psychiatric and substance abuse disorders^1^. Genetic factors may also play a role, with one study suggesting that higher common variant liability to schizophrenia, as indexed by polygenic scores (PGS), is associated with poorer response to antipsychotics after 12 weeks of treatment^2^. Similarly, both longitudinal and cross-sectional research has demonstrated an association between higher schizophrenia PGS and greater severity of negative and cognitive symptom dimensions^3–5^.

Beyond polygenic risk, variation in genes encoding proteins key to pharmacokinetic or pharmacodynamic processes (“pharmacogenes”) may also influence response to antipsychotic medication^6^. Pharmacogenomic star alleles describe single or multiple genetic markers that alter the function of proteins influencing drug response. The most widely researched pharmacogenes are within the cytochrome P450 (CYP) family, which are implicated in drug metabolism and are highly variable within and across populations. While drug metabolism pathways are often well-studied biochemically, the extent to which between-person variability in these pathways influences drug effectiveness is still unclear. **Figure 1** illustrates how a pharmacogenomic alteration in a key metabolic enzyme might affect response to a medication. While such effects are intuitive and form the basis of successful prospective trials^7^, as of June 2024, only 10 antipsychotics have regulator-approved guidelines with actionable pharmacogenomic markers in the PharmGKB database^8^. Reviews from expert consortia report a similar number of drugs where pharmacogenomic information might be of use in clinical settings^9^, though they also highlight the paucity of studies in the area.

**Figure 1.**
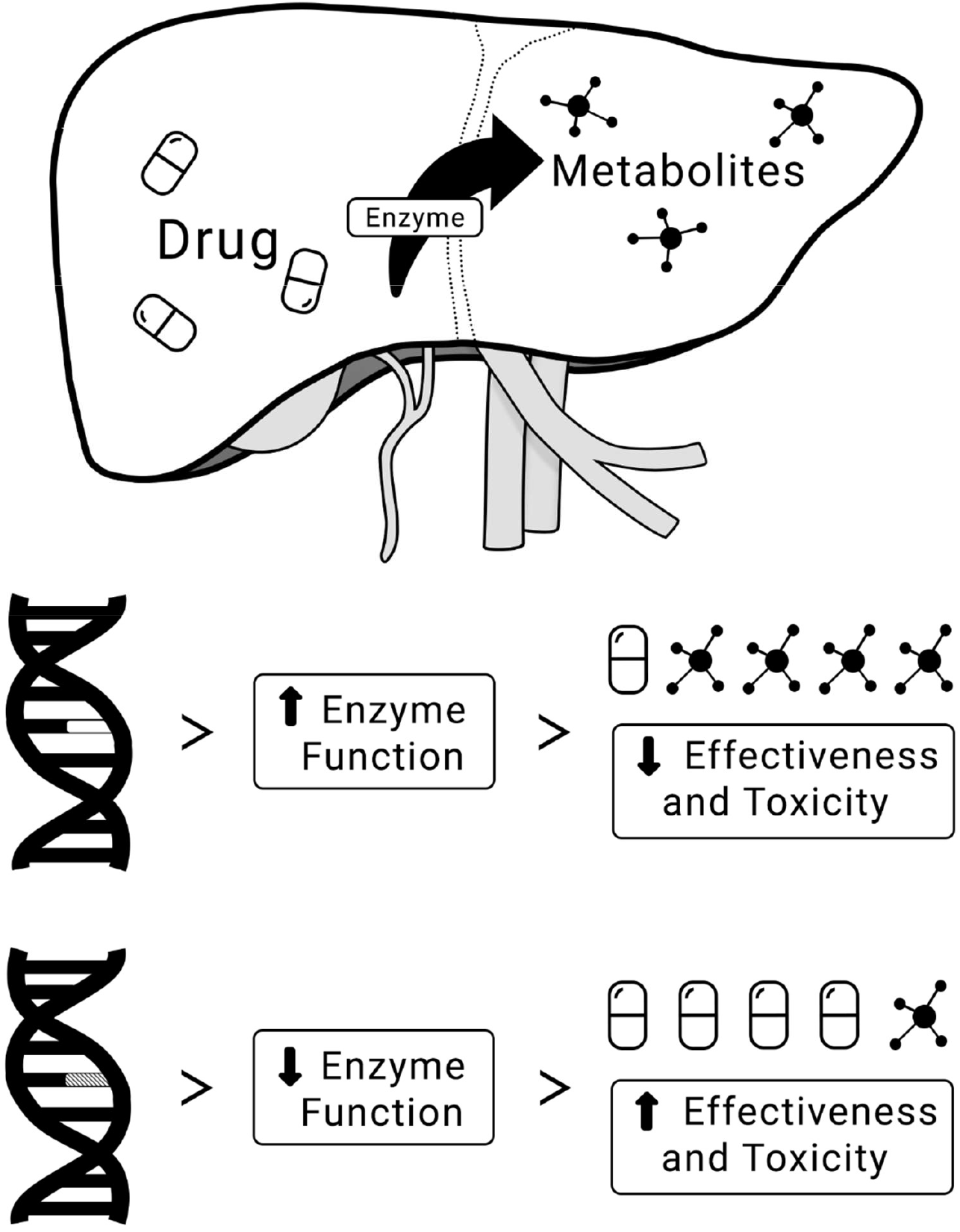
The majority of CYP-mediated drug metabolism occurs hepatically. Pharmacogenomic variation can result in increased, decreased, or no change to enzyme activity. Increased enzyme activity leads to faster metabolism, resulting in a lower drug concentration. This can be expected to lead to its reduced effectiveness but also a reduced risk for toxicity and adverse drug reactions. Conversely, variation leading to reduced enzyme activity results in slower metabolism of enzyme substrates. This can result in higher drug concentrations that might result in increased effectiveness but also increased risk of toxicity. The exact consequences of pharmacogenomic variation depend on the enzyme, drug, and metabolite(s) in question; for example, effects might vary if the metabolite has the potential to cause side effects or if the administered drug is a prodrug.

Establishing a pharmacogenomic marker as “actionable” requires an evaluation of its effects on phenotypes beyond drug metabolism and pharmacokinetics, and studies on clinical outcomes can be valuable in these decisions^9^. There is preliminary evidence that leveraging pharmacogenomic star alleles to infer enzyme activity may help predict schizophrenia symptom severity in those taking clozapine, the only evidence-based medication for treatment-resistant schizophrenia (TRS). Clozapine has a complex metabolic pathway but most of its first-pass bioconversion is driven by CYP1A2, with minor contributions of CYP2C19 and CYP2D6^10^. Two recent studies reported associations between schizophrenia symptom severity and genotype-predicted enzyme activity for CYP1A2^11^ and CYP2C19^12^ in people with TRS taking this medication. However, the effects reported were inconsistent between studies, and their interpretation is complicated by differences in the statistical methodology and symptom rating scales used.

The present study aims to investigate possible links between medication and pharmacogenomic variables with current schizophrenia symptom severity and cognitive ability in a UK-based sample of individuals treated with antipsychotic medication. Although cross-sectional, the inclusion of pharmacogenomic information alongside medication variables offers the potential to inform the causal direction of associations. We also reproduced previous pharmacogenomic analyses^11,12^ in a subsample restricted to patients prescribed clozapine. Our work follows recent calls to overcome “one size fits all” approaches in psychopharmacology by directly searching for predictors of clinical outcomes and pharmacodynamics^13,14^, as these are essential to advancing the field of precision psychiatry through effective patient stratification.

## Methods

### Participants

We included 585 individuals from the Cardiff COGnition in Schizophrenia (CardiffCOGS) cohort (**Supplementary Figure 1**). CardiffCOGS received approval from the South-East Wales Research Ethics Committee (07/WSE03/110) and all participants provided written informed consent. Participants were recruited from inpatient and community adult mental health services and voluntary services across the UK. Patients were also recruited from clozapine clinics; therefore, the cohort was enriched for those with treatment resistance (TRS; N = 37%). All participants completed a clinical research interview based on the Schedules for Clinical Assessment in Neuropsychiatry^15^ (SCAN) and provided a blood sample for genetic analyses. All participants met DSM-IV^16^ or ICD-10^17^ diagnoses for schizophrenia or schizoaffective disorder, depressed type based on the SCAN-based interview and clinical case note review. A full description of the cohort with additional details is provided elsewhere^4^.

### Phenotype Data

The severity of positive, negative, affective, and disorganised symptoms were assessed at the time of the interview and are referred to hereafter as “current” symptoms. Symptoms were rated using the Scale for the Assessment of Positive Symptoms^18^ (SAPS), the Scale for the Assessment of Negative Symptoms^19^ (SANS), and the Calgary Depression Scale for Schizophrenia^20^. Cognitive ability was assessed using the MATRICS Consensus Cognitive Battery^21^ (MCCB). Missing values for MATRICS variables were imputed in accordance with guidance from the MCCB handbook before being standardised against unaffected controls.

All participants were treated with antipsychotics at the time of the interview. Medication data were obtained via self-report and case note review. A binary medication adherence variable was created from an ordinal questionnaire response that was based on the Medication Adherence Rating Scale^22^ (see **Supplementary Materials**). Antipsychotic doses were converted to chlorpromazine equivalents using the *ChlorpromazineR* package^23^. Doses were converted based on international consensus values where available, and World Health Organisation Daily Defined Dose variables otherwise^24,25^. Where individuals reported taking multiple antipsychotics concurrently, each antipsychotic dose was converted and then summed to give the total chlorpromazine-equivalent dose. Participants (N = 88) were excluded when (i) no adherence information was available or when, for any antipsychotic reported: (ii) the drug name was not documented or (iii) no dose information was reported.

A binary index for the presence/absence of current anticholinergic drug use was created based on the current prescription of Procyclidine, Hyoscine, Benztropine, Benzhexol, or Pirenzepine. The Global Assessment Scale^26^ was used as a measure of current functioning.

### Genetic Data

Participants were genotyped with the Illumina HumanOmniExpressExome v8 or the Illumina HumanOmniExpress v12 (Illumina Inc, USA) at the Broad Institute of Harvard and MIT, MA, USA and deCODE Genetics, Reykjavík, Iceland. This procedure and the curation and harmonisation of data from different arrays have been described previously^27^. Combined genetic data for all samples were processed using the DRAGON-data quality-control pipeline “GenotypeQCtoHRC”^28^ with default parameters and imputed on the Michigan Imputation Server (see **Supplementary Materials**).

Pharmacogenomic markers for enzymes known to metabolise antipsychotic drugs (CYP1A2, CYP2C9, CYP2C19, CYP2D6, and CYP3A5) were called using the *run-chip-pipeline* command in PyPGx v0.20.0^29^ in Python v3.9.2^30^. Activity scores were derived from pharmacogenomic alleles as described in the **Supplementary Materials** and **Supplementary Table 1**.

PGS for schizophrenia^31^, intelligence^32^, and educational attainment^33^ were calculated using PRS-CS^34^ and PLINK v1.9^35^. SNP effect sizes (BETA/Odds Ratio) were used alongside standard errors to compute posterior effect sizes in PRS-CS. The 1000 Genomes Project phase 3 European Linkage Disequilibrium (LD) reference panel was used to account for linkage disequilibrium^36^. Additional parameters in PRS-CS included 10,000 burn-in iterations, 25,000 Markov chain Monte Carlo iterations, and a global shrinkage parameter phi of 1 for schizophrenia or “auto” for intelligence and education attainment. Effect sizes generated in PRS-CS were passed to PLINK v1.9 for scoring. PGS were based on custom GWAS datasets from which all participants of the current study had been excluded.

To account for population stratification, a subset of common SNPs with high imputation quality (MAF > 0.05, INFO > 0.9), and low levels of LD (r^2^ < 0.2) was selected to calculate principal components using the principal component analysis function in PLINK v2^35^.

### Statistical Analysis

Data were analysed using R v4.4.0 in R Studio 2024.04.0 Build 735^37^. Confirmatory Factor Analysis (CFA) was used to derive a latent factor model for current symptom severity in schizophrenia based on other latent models^4,5,38,39^. Variables contributing to the model were global symptom measures from the SAPS and SANS, the self-report depressed mood and suicidal ideation and acts items from the Calgary Depression Scale for Schizophrenia, and the domain scores (excluding social cognition) from MATRICS. Ratings from SAPS, SANS, and Calgary Depression Scale for Schizophrenia were ordinal (0 – 5, SAPS and SANS; 0 – 3, Calgary Depression Scale for Schizophrenia). MATRICS variables were continuous. *lavaan* was used to fit the CFA, using default settings^40^. Dimension scores based on the best fit CFA model were calculated for each participant. Model fit was guided by the Comparative Fit Index, Root Mean Square Error of Approximation, and the Standardised Root Mean Squared Residual. We would expect that current phenotype severity, indexed by latent variable scores, would be associated with functional impairments. Therefore, as an in-sample validity check of our derived variables, we regressed each factor against the Global Assessment Scale.

Linear regression models were used to test for associations between medication variables and symptom dimensions. Medication variables included daily chlorpromazine-equivalent antipsychotic dose, clozapine use, and anticholinergic use. All continuous variables were standardised, and all models controlled for schizophrenia PGS, medication adherence, age, sex, and the first 5 genetic principal components. Finally, intelligence and educational attainment PGS were used as an estimate for premorbid intelligence in the analyses investigating the cognitive ability dimension. We chose these PGS as together they are the strongest genetic predictor of intelligence in previous research^41,42^, and neither variable was associated with chlorpromazine-equivalent antipsychotic dose in our total sample. In contrast, premorbid intelligence estimated from the National Adult Reading Test (NART)^43^ was associated with dose indicating the possibility that antipsychotic dose influenced performance on the NART or vice versa. Nevertheless, an alternative analysis using premorbid intelligence as estimated by the NART is provided in the **Supplementary Materials**.

The model was extended by including activity scores for CYP1A2, CYP2C9, CYP2C19, CYP2D6, and CYP3A5 to test their potential to mediate or moderate any associations in the total sample, and in the subgroup of participants prescribed clozapine (N = 215) separately. Note that analyses of the effects of the pharmacogenomic variables are adjusted for dose, clozapine use, and anticholinergic use. Finally, we performed a sensitivity analysis to test whether our pharmacogenomic models changed after controlling for concomitant medication or lifestyle factors that may influence enzyme function (“phenoconversion”^44^) as described in the **Supplementary Materials**.

This study followed the STrengthening the REporting of Genetic Association Studies reporting recommendations^45^ (STREGA), an extension of the STROBE Statement^46^. The checklist can be found in the **Supplementary Materials**.

## Results

We included 585 participants with schizophrenia or schizoaffective disorder, depressed type (205 [35%] female; mean [SD] age of 43.5 [11.7] years). At the time of the interview, participants were prescribed at least one of 16 different antipsychotics, administered orally or by long-acting injection. Clozapine (36.8%) and olanzapine (17.6%) were the most frequently prescribed antipsychotics.

Descriptive statistics **(Supplementary Table 2)**, the frequency of antipsychotics reported **(Supplementary Table 3)**, and the allele frequency of pharmacogenomic star alleles **(Supplementary Table 4)** are detailed in the **Supplementary Materials**.

### Confirmatory Factor Analysis of Schizophrenia Phenotypes

Four factor structures were fit to determine which best represented the data as described in **Supplementary Table 5**. The model with the best fit (Comparative Fit Index = 0.997, Root Mean Square Error of Approximation = 0.019, Standardised Root Mean Square Residual = 0.043) had five dimensions related to positive symptoms, negative symptoms of diminished expressivity, negative symptoms of reduced motivation and pleasure, depression and suicide, and cognitive ability (**Figure 2**). The derived factors were all significantly associated with the Global Assessment Scale in the expected direction (**Supplementary Materials**).

**Figure 2.**
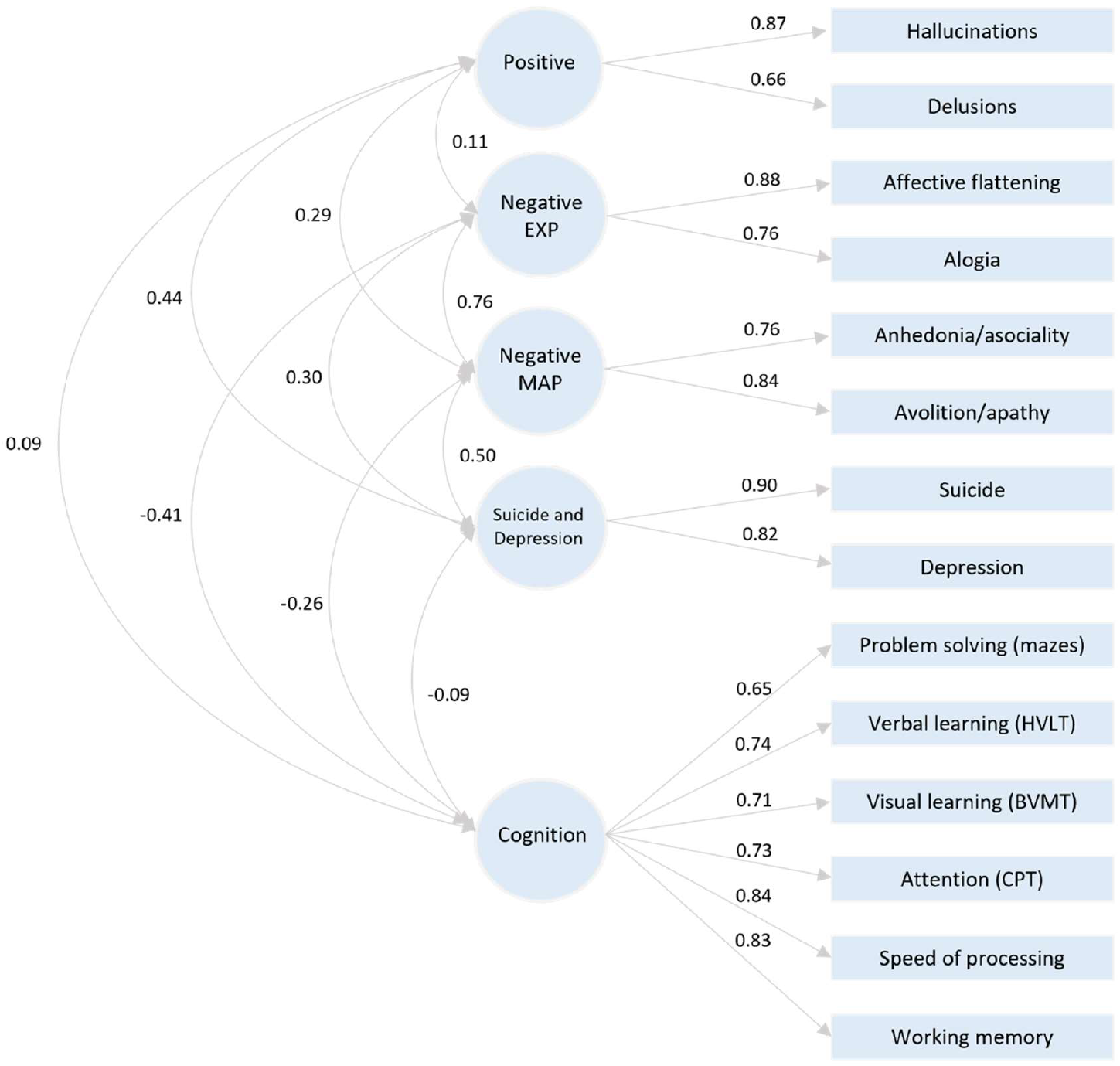
Factor structure from the confirmatory factor analysis of current schizophrenia phenotypes within the CardiffCOGS sample. Standardised loadings between latent factors (circles) and contributing phenotypes (rectangles) are reported on straight lines. Curved lines represent correlations between latent factors. EXP, Diminished Expressivity; MAP, Reduced Motivation and Pleasure; HVLT, Hopkins Visual Learning Test; BVMT, Brief Visuospatial Memory Test-Revised; CPT, Continuous Performance Test.

### Associations between Phenotype Dimensions and Medication Variables

Only associations that passed the 5% False Discovery Rate (FDR) threshold were considered significant. Effect sizes were standardised and shown alongside uncorrected *p* values. The results of association tests between phenotype dimensions and medication variables are shown in **Table 1**. Extended versions of this (and other tables) that include associations between further covariates and phenotype dimensions are given in **Supplementary Tables 6 – 8**.

**Table 1.**
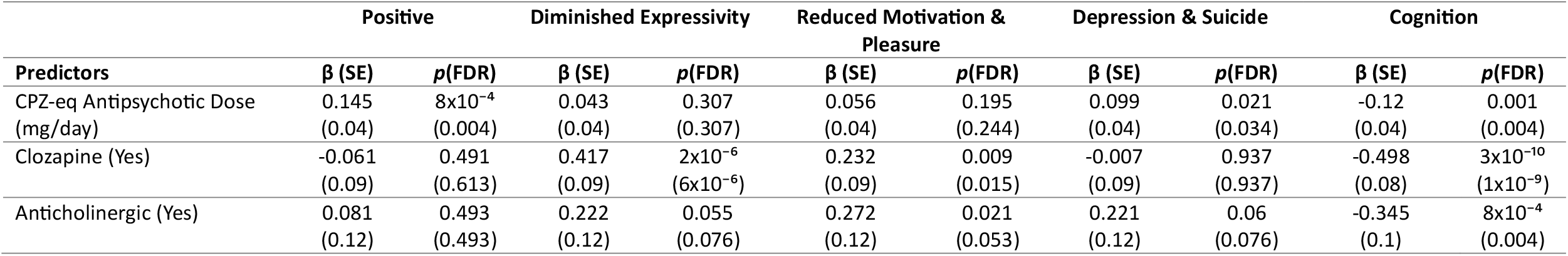
Associations between medication variables and severity of the five examined schizophrenia phenotype dimensions. Associations between medication variables with schizophrenia symptom and cognitive ability dimensions in CardiffCOGS. Five separate models were fit, each with severity scores for a schizophrenia phenotype as the outcome variable. Variables of interest included the medication variables, chlorpromazine-equivalent daily antipsychotic dose, clozapine use, and anticholinergic use. All models controlled for medication adherence, schizophrenia PGS, age, sex, and the first five genetic principal components. Intelligence PGS and Educational Attainment PGS were included as additional covariates in the model where the cognitive ability dimension is the outcome. Standardised regression estimates are reported. CPZ-eq = chlorpromazine-equivalent; SE = Standard Error; PGS = Polygenic Score.

Higher scores on the positive dimension were associated with higher chlorpromazine-equivalent antipsychotic dose (β = 0.145; 95%CI, 0.06 to 0.23; *p* = 8×10^−4^). Higher scores on the diminished expressivity dimension were associated with clozapine use (β = 0.417; 95%CI, 0.25 to 0.59; *p* = 2×10^−6^). Higher scores on the reduced motivation and pleasure dimension were also associated with clozapine use (β = 0.232; 95%CI, 0.06 to 0.41; *p* = 0.009). Higher scores on the suicide and depression dimension were associated with higher chlorpromazine-equivalent antipsychotic dose (β = 0.099; 95%CI, 0.02 to 0.18; *p* = 0.021).

Lower scores on the cognition dimension, indicating poorer cognitive ability, were associated with higher chlorpromazine-equivalent daily antipsychotic dose (β = -0.12; 95%CI, -0.19 to -0.05; *p* = 0.001), clozapine use (β = -0.498; 95%CI, -0.65 to -0.35; *p* = 3×10^−10^), and anticholinergic use (β = - 0.345; 95%CI, -0.55 to -0.14; *p* = 8×10^−4^).

### Pharmacogenomic Associations with the Phenotype Dimensions

All pharmacogenomic associations were adjusted for medication variables. The addition of pharmacogenomic variables into the models did not substantially influence the associations between the medication variables and schizophrenia phenotype dimensions described above. While some attenuation of *p* values was observed, there was no change to the overall pattern of results, with previously significant associations remaining so.

In the full sample, we found several associations in which lower symptom dimension scores, and so less severe symptoms, were associated with faster genotype-inferred enzyme activity. Lower scores on the positive symptom dimension were associated with higher CYP2C19 activity scores (β = -0.108; 95%CI, -0.19 to -0.03; *p* = 0.009). Lower scores on the diminished expressivity dimension were associated with higher CYP3A5 activity score (β = -0.113; 95%CI, -0.19 to -0.03; *p* = 0.007). Similarly, we also found an association between lower scores on the reduced motivation and pleasure dimension and higher CYP3A5 activity score (β = -0.106; 95%CI, -0.19 to -0.02; *p* = 0.012). There were no pharmacogenomic associations with either the suicide and depression or the cognitive ability dimension (**Table 2**).

**Table 2.**
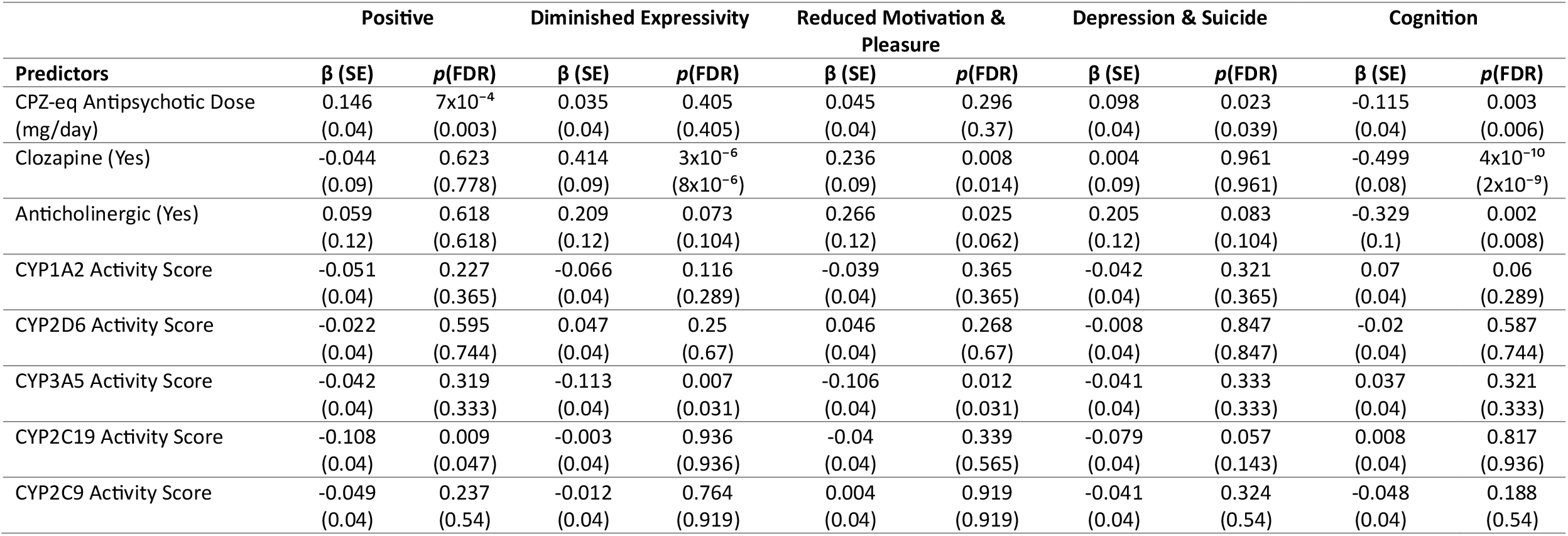
Associations between medication and pharmacogenomic variables and severity of the five examined schizophrenia phenotype dimensions. Associations between medication and pharmacogenomic variables with schizophrenia symptom and cognitive ability dimensions in CardiffCOGS. Five separate models were fit, each with severity scores for a schizophrenia phenotype as the outcome variable. Variables of interest included the medication variables (i.e., chlorpromazine-equivalent daily antipsychotic dose, clozapine use, and anticholinergic use) and pharmacogenomic variables (i.e., the genetics-inferred enzyme activity scores). All models controlled for medication adherence, schizophrenia PGS, age, sex, and the first five genetic principal components. Intelligence PGS and Educational Attainment PGS were included as additional covariates in the model where the cognitive ability dimension is the outcome. Standardised regression estimates are reported. CPZ-eq = chlorpromazine-equivalent; SE = Standard Error; PGS = Polygenic Score.

Within the subgroup of participants taking clozapine, higher CYP1A2 activity score was associated with higher cognitive ability (β = 0.17; 95%CI, 0.05 to 0.29; *p* = 0.005). No other pharmacogenomic variables were significantly associated with schizophrenia phenotype severity after correcting for multiple comparisons in this clozapine taking group (**Table 3**), although higher dose of antipsychotic was nominally associated with lower cognitive ability (β = -0.147; 95%CI, -0.27 to -0.02; *p* = 0.02).

**Table 3.**
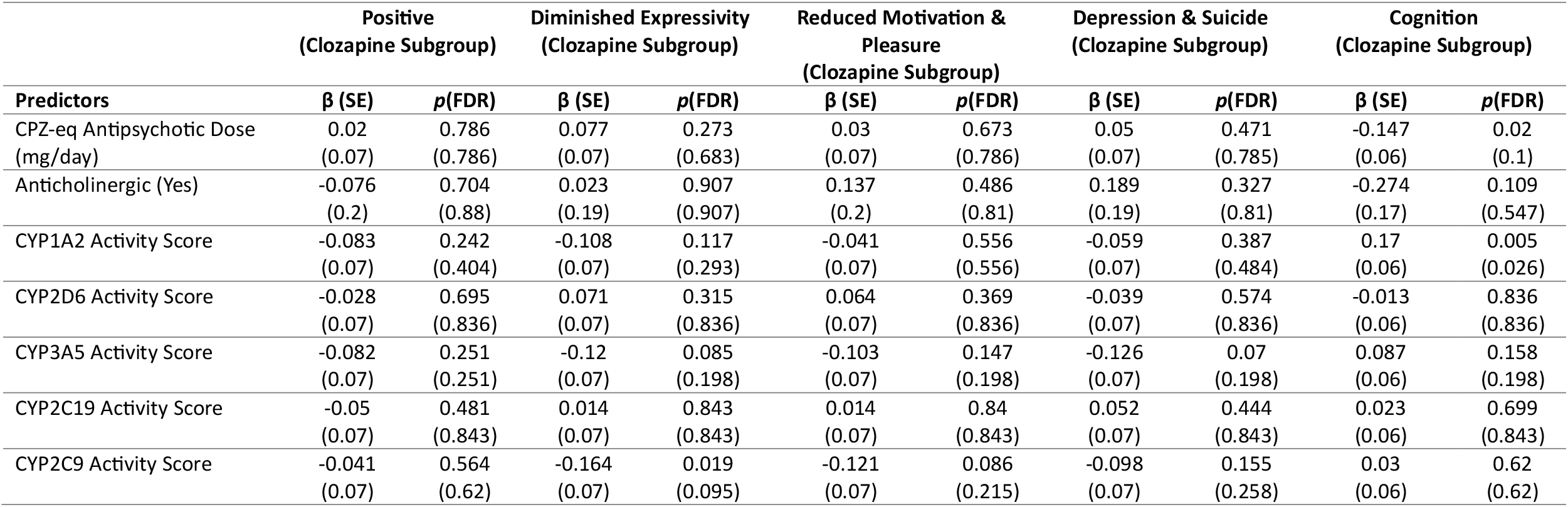
Associations between medication and pharmacogenomic variables and severity of the five examined schizophrenia phenotype dimensions in the subgroup of participants prescribed clozapine. Associations between medication and pharmacogenomic variables with schizophrenia symptom and cognitive ability dimensions in the subgroup of CardiffCOGS participants prescribed clozapine. Five separate models were fit, each with severity scores for a schizophrenia phenotype as the outcome variable. Variables of interest included the medication variables (i.e., chlorpromazine-equivalent daily antipsychotic dose, clozapine use, and anticholinergic use) and pharmacogenomic variables (i.e., the genetics-inferred enzyme activity scores). All models controlled for medication adherence, schizophrenia PGS, age, sex, and the first five genetic principal components. Intelligence PGS and Educational Attainment PGS were included as additional covariates in the model where the cognitive ability dimension is the outcome. Standardised regression estimates are reported. CPZ-eq = chlorpromazine-equivalent; SE = Standard Error; PGS = Polygenic Score.

As recommended in the STREGA guidelines, we provide unadjusted estimates for associations between pharmacogenomic variables and symptom dimensions (**Supplementary Tables 9 & 10)**. Sensitivity analyses in which we controlled for medication that may lead to phenoconversion did not lead to different results (**Supplementary Tables 11 & 12**). However, inclusion of an interaction term between CYP1A2 and smoking status in our model attenuated the association between CYP1A2 activity score and the cognitive dimension (**Supplementary Table 13)**. This suggests that this pharmacogenomic association may be partly explained by patient lifestyle factors. However, our sensitivity analysis is limited by incomplete smoking information in the sample; thus, diminished statistical power may also account for this weakened association.

## Discussion

This cross-sectional study investigated medication and pharmacogenomic correlates of current phenotype severity in people with schizophrenia or schizoaffective disorder depressed-type. We found that higher daily chlorpromazine-equivalent antipsychotic dose was associated with more severe scores on both the positive symptom and the suicide and depression dimensions, while clozapine use was associated with worse scores on the two negative symptom dimensions (i.e., diminished expressivity, reduced motivation and pleasure). We also found that chlorpromazine-equivalent daily antipsychotic dose, clozapine use, and anticholinergic use were all associated with lower scores on the cognitive dimension, indicating poorer cognitive ability.

Some of the associations we have observed between drug dosage and symptomatology are likely to reflect influences of the latter on prescribing patterns. Higher doses of antipsychotic medication are likely to be prescribed to help manage more severe or incompletely resolved positive symptoms, while negative symptoms do not generally respond well to typical antipsychotics and may trigger the prescription of clozapine which does have efficacy in this domain^47^.

Our findings that antipsychotic dose, clozapine use, and anticholinergic use were associated with poorer cognition are consistent with other research^48–53^. Moreover, our effect sizes were, in some instances, substantial, with our strongest association indicating that scores on the cognitive ability dimension are nearly 0.5 standard deviations lower in patients prescribed clozapine. These findings could be plausibly related, in part, to causal effects of drug treatment. However, interpreting cross-sectional findings relating to cognition and medication use is challenging due to the possibility of reverse causation whereby patients with cognitive impairment could be more likely to receive higher doses of medication. The situation with respect to clozapine may be even more complex because a prescription of clozapine usually necessitates a diagnosis of TRS, which is itself associated with greater cognitive impairment than treatment-responsive schizophrenia^54–56^. Nevertheless, the associations observed between both antipsychotic dose and clozapine use with poorer cognition were significant after controlling for premorbid intelligence as indexed by PGS for intelligence and educational attainment. These medication associations mostly replicate in the alternative analysis described in the **Supplementary Materials** using NART to estimate premorbid intelligence, albeit with weaker signals. In all, our analyses tentatively support the interpretation that the association may be causally related to medication use, instead of reflecting the confounding effects of lower premorbid cognitive ability in those who subsequently receive clozapine or higher doses of antipsychotics.

Our combined analysis of pharmacogenomic and medication variables offers a potential means of drawing causal inferences in regard to the observed associations between drug treatment and cognition. CYP1A2 is the main enzyme responsible for clozapine first-pass metabolism^57,58^. Higher enzyme activity can be expected to result in lower clozapine bioavailability for a given dose. Under a causal model, this should lead to fewer clozapine-induced cognitive effects. We indeed observed this in the association between higher cognitive ability and faster genotype-inferred CYP1A2 activity in people prescribed clozapine (**Table 3**), which points to the possibility that any negative impact of clozapine in cognition could be ameliorated in those with faster clozapine metabolism. In parallel, a nominally significant association also exists between higher cognitive ability and lower dose in those prescribed clozapine. In the context of all these results, we remark the use of pharmacogenetic information to phenotype enzyme activity and thus drug metabolism, highlighting individuals that could be particularly vulnerable to dose-dependent adverse drug reactions (ADRs). We also note that while this research takes advantage of such a phenotyping approach, the atypical instances of clozapine metabolism that would characterise these individuals in clinical scenarios can already be detected by therapeutic drug monitoring. For this reason, we echo the recent recommendations for therapeutic drug monitoring procedures to be more readily adopted as a routine part of the clinical management of those on clozapine^59^.

We also reported an inverse association between prescription of anticholinergic medication and cognition. Some antipsychotics cause extra-pyramidal side effects, generally as a dose-dependent ADR^60,61^. Extra pyramidal side effects are managed with anticholinergic medication, and although not recommended, these drugs may be prescribed prophylactically, pre-empting the onset of these ADRs^62,63^. The association between anticholinergic prescription and cognition is independent of daily antipsychotic dose. Therefore, this likely reflects a distinct contribution of anticholinergics on cognition, as opposed to capturing our reported dose-cognition association given the higher rates of anticholinergic prescription amongst people on high antipsychotic doses^64^.

We observed several associations between faster genotype-inferred enzyme activity with lower schizophrenia symptom severity. Increased CYP2C19 activity was associated with lower severity of positive symptoms. Associations between CYP2C19 with both schizophrenia severity^12,65^ and symptom improvement^66^ have been reported although those studies included only people taking clozapine and employed a single, general measure of symptom severity. In the absence of longitudinal symptom scores, we cannot investigate the rather counter-intuitive hypothesis suggested by our data that greater antipsychotic clearance, indexed by CYP2C19 activity, might lead to increased drug effectiveness. CYP2C19 is thought to play a minor part within the clozapine metabolic pathway^67^ but its role in antipsychotic metabolism more widely is poorly documented^68^. This is an avenue for future studies to follow.

Increased genetically inferred CYP3A5 activity was associated with lower scores on both the negative symptom dimensions. While associations with the two domains increases confidence in the findings, caution is warranted given that these are moderately correlated (see **Figure 2**), and we did not find orthogonal evidence that either of these dimensions were associated with antipsychotic dose. Thus, replication of these associations is required. CYP3A5 is relatively under-examined in antipsychotic research^69^ and its main genotype that leads to poor metabolism (*1/*3) is common worldwide but particularly in European populations. Indeed, it has been observed that major differences in genotype/phenotype distributions for this enzyme are mainly driven by the inclusion of African ancestries in studies^70,71^, as functional CYP3A5 alleles are more common in Africa. Populations from Asian countries also seem to have higher diversity of CYP3A5 alleles than Europe, though research in this continent is still scarce^72^. Future research aiming to investigate the relevance of CYP3A5 for outcomes in psychiatric populations should therefore aim to include more cohorts from admixed and non-European ancestral makeups.

Finally, we did not observe evidence for an association between CYP1A2 activity score and either the positive or negative schizophrenia symptom dimensions. A correlation between CYP1A2 activity and symptom severity has been reported^11^, and while several differences exist between the methods and sample employed in our own and the previous study, the observed trends in our data are not consistent with their findings. We also found no association between the CYP2C9 activity score and any of our phenotype severity scores.

The primary limitation of this study is its cross-sectional nature which means we are unable to assess drug treatment response, constraining our ability to make robust inferences about the causal directions for any of the associations. Second, our best fitting CFA model did not include a disorganised dimension, and therefore our study cannot address the possible relationships between this symptom dimension with medication and pharmacogenomic variables. Finally, although no individuals were excluded based on ancestry, 95% of the sample reported UK/European as their ethnic background. Cross-cultural differences permeate most aspects of psychiatry, with variation existing in diagnostic criteria, prognosis, therapeutic/prescribing practices, and more^73^. While ancestrally diverse samples are becoming more common, particularly in larger scale genetics studies, this hasn’t always been the case. Therefore, replication of this work across ancestrally diverse samples is required to determine whether these results are generalisable outside of White European backgrounds.

Overall, our findings indicate that poorer cognitive ability in individuals with schizophrenia was associated with the use of clozapine and anticholinergics, alongside high doses of antipsychotic medication. Cognition is an important predictor of schizophrenia functional outcomes^74^. Therefore, understanding the potentially multifaceted burden of these drugs could help clinicians minimise the likelihood of cognitive impairment and other poor outcomes during schizophrenia pharmacotherapy. Longitudinal studies, particularly randomised controlled trials, are required to fully understand the role that pharmacotherapy, especially clozapine and anticholinergics, has on cognitive outcomes for those with schizophrenia. However, given the lack of evidence-based guidelines for optimising antipsychotic doses in the maintenance phase of treatment^75^, addressing potential cognitive impacts might be an actionable target for future interventional research. For example, if information from prescription records (e.g. drug classes and doses) could contribute to identifying patients at high risk of reduced cognition, they could be prioritised for closer monitoring and/or mitigations (e.g., cognitive remediation therapy or avoiding prophylactic anticholinergics).

We also identify associations between the increased activity of certain CYP enzymes and the reduced severity of positive, negative, and cognitive symptoms. Except for CYP1A2 in patients taking clozapine, which we discuss earlier, the mechanisms by which variation within these pharmacogenes could influence schizophrenia severity are unclear. While our results implicate pharmacogenomic variation in antipsychotic pharmacodynamics, an area of psychopharmacology where robust predictors are particularly scarce, further validation of our findings in larger, more diverse samples is required before charting a course from this basic evidence towards improved strategies for patient support and care.

## Data Availability

Code for reproducing all the main analyses in R is available online at https://locksk.github.io/cogs-symptoms/). Additional scripts for calling pharmacogenomic star alleles and calculating polygenic scores is available online at https://github.com/locksk/symptoms-medication-pgx. To comply with the ethical and regulatory framework of the CardiffCOGS project, access to individual-level data requires a collaboration agreement with Cardiff University. Requests to access deidentified datasets, data dictionaries, and other summaries from the CardiffCOGS project should be directed to James Walters (waltersjt{at}cardiff.ac.uk)

https://github.com/locksk/symptoms-medication-pgx

## Notes

### Competing Interest Statement

MJO, MCOD, and JTRW are supported by a collaborative research grant from Takeda Pharmaceuticals Ltd. for a project unrelated to work presented here. AFP, MJO, MCOD, and JTRW also reported receiving grants from Akrivia Health for a project unrelated to this submission. Takeda and Akrivia Health played no part in the conception, design, implementation, or interpretation of this study.

### Funding Statement

This project was supported by a Medical Research Council Programme Grant (MR/Y004094/1) and The National Centre for Mental Health, funded by the Welsh Government through Health and Care Research Wales. SKL was funded by a PhD studentship from Mental Health Research UK (MHRUK). DBK, JTRW, MCOD and AFP were supported by the European Union's Horizon 2020 research and innovation programme under grant agreement 964874.

