## Supplemental Materials for "Medication and pharmacogenomic effects on cross-sectional symptom severity and cognitive ability in schizophrenia"

### Supplementary Methods

#### Adherence Questionnaire Item

Adherence to current medication was measured on an ordinal scale as described below. Participants were rated according to the most appropriate statement:

0) Doesn't take medication.
1) Takes medication only rarely (1-2/week).
2) Misses over 50% doses.
3) Misses doses every week.
4) Rarely misses doses.
5) Medication supervised and takes as prescribed.

Responses were dichotomised due to low numbers of ratings in certain categories. Participants who missed doses every week, or more frequently (0-3) were assigned as non-adherent. Those who reported rarely missed doses, or were medication supervised (4-5) were assigned as adherent.

#### Quality control of Genetic Data

Pre-imputation quality control (QC) was performed as follows; SNPs were excluded due to call rates < 0.95, MAF < 0.01, or a Hardy-Weinberg Equilibrium mid-$p < 10^{-6}$. Individuals were excluded with genotyping coverage rates < 0.95 or potential errors in a PLINK "sex check" analysis (i.e., ambiguous genetic sex, or mismatch between self-reported and genetic sex).

The Michigan Imputation server[1] and Minimac-4 were utilised for imputation, using the Haplotype Reference Consortium v1.1 as the reference panel. Pre-imputation statistical phasing was carried out in the Michigan built-in service using Eagle 2.4[2]. Post imputation, SNPs were removed when they had a hard-call genotype probability threshold < 0.9, $R^2 < 0.9$, MAF < 0.01, and genotyping rates < 0.95. Relatedness was assessed through the PC-Relate algorithm[3] and a cutoff of $\pi > 0.4$ was used to remove related individuals[4].

For pharmacogenomic allele calling we retained any monomorphic variants in the dataset and used a more relaxed imputation quality threshold ($R^2 < 0.7$; Hard-call genotype probability < 0.8). This allowed us to retain a greater number of pharmacogenomic informative SNPs to increase the allele calling accuracy, while still being stringent enough to avoid imputation errors in common genetic variants[5,6]

#### Pharmacogenomic Activity Score Mapping

PyPGx v0.20.0[7] predicts enzyme metabolism phenotypes in two ways, through activity scores or diplotype-phenotype mapping. Despite this, it does not explicitly report activity scores in its output. Where available, activity scores were mapped to pharmacogenomic alleles called using the allele table within the PyPGx package. This was feasible for CYP2D6 and CYP2C9. For the remaining enzymes, the mapping of pharmacogenomic alleles to activity scores was based on existing literature or inferred from PharmGKB Allele Functionality Reference Tables (see **Supplementary Table 1**).

For CYP1A2, PharmGKB does not yet provide an allele functionality table. Thus, activity score assignment was based on consensus values from the literature[8,9]. No guidance was available for *1K, therefore it was assigned the same activity score as *1C, also a decreased function allele. Where *CYP1A2**1F/1C was called on a single haplotype, we assigned an activity score of 1 based on past work[10].

#### Accounting for phenoconversion

Phenoconversion describes the gap between genetically inferred enzymatic status and actual enzyme activity[11]. Many different factors can influence enzyme activity, and in particular CYP enzymes may be induced or inhibited by a host of variables including concomitant medication, lifestyle factors, diet, and inflammation[12].

We observed associations between three pharmacogenomic activity scores and schizophrenia phenotype dimensions. Based on the Food and Drug Administration's (FDA) table of CYP Enzyme- and Transporter System-Based Clinical Substrates, Inhibitors, or Inducers[13], we identified three drugs within the CardiffCOGS sample that could influence the activity of these enzymes. These drugs include the CYP2C19 strong inhibitor fluoxetine (N = 34), the CYP3A5 strong inducer carbamazepine (N = 5), and the CYP1A2 moderate inducer cigarette smoking (N = 305). We are however unable to account for other drugs and factors not reported in our study (e.g., oral contraceptives, dietary factors, disease, and inflammation) that may further influence enzyme activity[11,12].

We accounted for phenoconversion using two methods, (i) statistically through an interaction term between the enzyme activity score and a binary variable representing use of the relevant drug, and (ii) traditionally by replacing standard activity scores with phenoconversion-corrected activity scores. Phenoconversion-corrected activity scores were calculated based on work by Lesche and colleagues[9]. Scores were multiplied by 1.5 in the presence of an inducer, and by 0 when a strong inhibitor was present. Models were fit only where a pharmacogenomic variable was associated with a phenotype dimension in our previous analyses, for example:

i) positive ~ CYP2C19 activity score * fluoxetine use + …
ii) positive ~ CYP2C19 phenoconversion-corrected activity score + …

Medication variables and control covariates were included within the models as in our main analyses, and each model only included one pharmacogenomic variable.

#### Estimating premorbid intelligence with NART

Associations between cognitive ability and medication variables could reflect a causal effect of medication use on cognition, or equally that people with lower cognitive ability are more likely to be prescribed higher doses or different medications to those with higher cognitive ability. Therefore, we aimed to control for a measure of premorbid intelligence in the regression models. For analysis, NART was converted to predicted WAIS-R full scale IQ via the recommended transformation IQ=130.6-1.24*NART error score. We compared two measures that estimate premorbid intelligence (the NART and intelligence and education PRS) by investigating their association with current cognition and chlorpromazine-equivalent antipsychotic dose in the total sample:

i) cognition ~ NART score + Age + Sex
ii) cognition ~ intelligence PGS + education attainment PGS + Age + Sex + PC1:5
iii) daily dose ~ NART score + Age + Sex
iv) daily dose ~ intelligence PGS + education attainment PGS + Age + Sex + PC1:5

#### Supplementary Results

##### Associations with the Global Assessment Scale

Four factor structures were fit to determine which best represented the data as described in **Supplementary Table 4**. Scores for all latent variables were significantly associated with current Global Assessment Scale ratings in models that controlled for age and sex. In all instances, increased symptom severity was associated with reduced scores on the Global Assessment Scale. This manifested as inverse associations between the positive (β = -0.54; 95%CI, -0.62 to -0.46; $p = 6x10^{-37}$), diminished expressivity (β = -0.416; 95%CI, -0.5 to -0.33; $p = 2x10^{-21}$), reduced motivation and pleasure (β = -0.544; 95%CI, -0.62 to -0.47; $p = 4x10^{-39}$), and depression/suicide dimensions (β = -0.543; 95%CI, -0.62 to -0.47; $p = 3x10^{-37}$) with Global Assessment Scale scores. The cognition dimension was positively associated with GAS (β = 0.174; 95%CI, 0.09 to 0.26; $p = 3x10^{-5}$), with higher ratings across the Global Assessment Scale and the cognition dimension both reflecting improved functioning.

##### Sensitivity Analysis

*CYP2C19 Phenoconversion*

There was no evidence of a significant interaction between the CYP2C19 activity score and fluoxetine use (β = -0.123; 95%CI, -0.41 to 0.17; *p* = 0.41). However, CYP2C19 activity remained significantly associated with the positive symptom dimension (β = -0.09; 95%CI, -0.17 to -0.01; *p* = 0.037). Fluoxetine use was also inversely associated with positive dimension scores (β = -0.363; 95%CI, -0.71 to -0.02; *p* = 0.04). A phenoconversion-corrected CYP2C19 activity score was not associated with the positive symptom dimension (β = -0.022; 95%CI, -0.1 to 0.06; *p* = 0.6). Full results are reported in **Supplementary Table 11**.

*CYP3A5 Phenoconversion*

The number of individuals taking carbamazepine was very low (N = 5); therefore, we were unable to model an interaction between CYP3A5 with carbamazepine use. Equally, we did not calculate phenoconversion-corrected activity scores as all carbamazepine users had null function alleles – thus, their activity scores would have been unchanged (i.e., 0 x 1.5 = 0). The associations between CYP3A5 activity remained with both the diminished expressivity (β = -0.106; 95%CI, -0.19 to -0.02; *p* = 0.01) and reduced motivation and pleasure dimensions (β = -0.1; 95%CI, -0.18 to -0.02; *p* = 0.018) after accounting for carbamazepine use through a covariate, which was itself not associated with either symptom dimension. Full results are available in **Supplementary Table 12**.

*CYP1A2 Phenoconversion*

We saw no significant interaction between the CYP1A2 activity score and cigarette use (β = -0.018; 95%CI, -0.2 to 0.16; *p* = 0.85). Cigarette smoking was inversely associated with cognition (β = -0.29; 95%CI, -0.54 to -0.04; *p* = 0.026); however, the association between CYP1A2 activity and cognition weakened (β = 0.19; 95%CI, -0.01 to 0.39; *p* = 0.06). Finally, we observed no significant association between the phenoconversion-corrected CYP1A2 activity scores with cognition (β = -0.022; 95%CI, -0.15 to 0.11; *p* = 0.74). Full results are in **Supplementary Table 13**.

The *p* value for the association between CYP1A2 activity score and cognition became larger after controlling for smoking status, suggesting that part of the variance in cognition explained by CYP1A2 is due to the inductive effect of cigarette smoking. However, information regarding patient smoking status was absent for just under 10% of the original subgroup (N = 18). The attenuation of the CYP1A2-cognition association could be explained in part by this loss of statistical power. We note that the negative association between cigarette smoking, and cognition is somewhat contrary to expectations. Cigarette smoke is a known CYP1A2 inducer, binding with aryl hydrocarbon receptors and resulting in increased enzyme activity[14]. However, chronic smoking is also robustly associated with cognitive impairment[15]. Therefore, smoking may be exerting dual, contradictory effects on cognition in our sample; a larger, inverse direct effect, and a more modest indirect, positive effect through induction of CYP1A2 activity.

#### Exploring Measures of Premorbid Intelligence

*Comparison of premorbid intelligence measures*

Premorbid intelligence as estimated by the NART had a stronger association with the cognitive ability dimension (β = 0.517; 95%CI, 0.45 to 0.58; p = $5x10^{-47}$), compared to the intelligence PGS (β = 0.097; 95%CI, 0.01 to 0.19; p = 0.034) and educational attainment PGS (β = 0.115; 95%CI, 0.03 to 0.2; p = 0.012).

However, we also found that premorbid intelligence as estimated by the NART was inversely associated with chlorpromazine-equivalent antipsychotic dose (β = -0.131; 95%CI, -0.21 to -0.05; p = 0.002) whereas there was no such association with dose for either intelligence PGS (β = -0.034; 95%CI, -0.13 to 0.06; p = 0.495) or educational attainment PGS (β = -0.016; 95%CI, -0.11 to 0.08; p = 0.75).

The results could indicate that participants with lower premorbid intelligence scores from the NART were prescribed higher antipsychotic doses. An alternative interpretation is that performance on the NART was affected by the participant's antipsychotic dose (i.e. sedation effects) and therefore, might not fully reflect pre-morbid intelligence in this sample[16]. As we were not able to distinguish between these potential confounding effects, we selected to use both intelligence and educational PGS as the estimates of premorbid intelligence in our primary analyses. However, we have conducted secondary analyses controlling for the NART, shown below.

*Alternative analyses controlling for premorbid intelligence as estimated by the NART*

This secondary analysis controls for premorbid intelligence as estimated by the NART, instead of PGS for intelligence and educational attainment, in analyses where the cognitive ability dimension is the outcome. Results are presented in **Supplementary Table 14.** NART data was not available for all participants in the full sample (N = 27) and those prescribed clozapine (N = 13); hence, these analyses have a lower sample size than those reported in the main text.

The associations between antipsychotic dose, clozapine use, and anticholinergic use with cognitive ability remained significant when including NART in the models, despite an attenuation in the strength of effect sizes and *p*-values.  However, in the pharmacogenomic model including only patients prescribed clozapine, covarying for NART reduced the effect size for the associations of cognitive ability with anticholinergic use and CYP1A2 activity score, rendering their *p*-values non-significant.

#### Supplementary Figures

Supplementary Figure 1. Ascertainment of sample size.

| | **N** |
|---|---|
| Total CardiffCOGS sample | 1310 |
| Genetic information available | 986 |
| Schizophrenia or Schizoaffective disorder depressed-type diagnosis | 747 |
| Medication information available | 620 |
| Symptoms/Cognition measures available | 587 |
| Not included in CLOZUK2/3 | 585 |
| Final Sample | 585 |

Supplementary Figure 1. Diagram showing participant exclusion based on several criteria and ascertainment of the final sample.

#### Supplementary Tables

##### Supplementary Table 1. CYP Pharmacogenomic star alleles and their respective activity scores.

| Enzyme | Allele | Function | Activity Score | Source |
|---|---|---|---|---|
| CYP1A2 | 1A | Normal | 1 | Lesche et al., 2019 |
| CYP1A2 | 1C | Decreased | 0.5 | Saiz-Rodriguez et al., 2019 |
| CYP1A2 | 1F | Increased | 1.5 | Lesche et al., 2019 |
| CYP1A2 | 1K | Decreased | 0.5 | - |
| CYP2D6 | 1 | Normal | 1 | pypgx |
| CYP2D6 | 2 | Normal | 1 | pypgx |
| CYP2D6 | 4 | Null | 0 | pypgx |
| CYP2D6 | 10 | Decreased | 0.25 | pypgx |
| CYP2D6 | 17 | Decreased | 0.5 | pypgx |
| CYP2D6 | 28 | Uncertain | N/A | pypgx |
| CYP2D6 | 35 | Normal | 1 | pypgx |
| CYP2D6 | 41 | Decreased | 0.5 | pypgx |
| CYP2D6 | 59 | Decreased | 0.5 | pypgx |
| CYP2D6 | 117 | Uncertain | N/A | pypgx |
| CYP3A5 | 1 | Normal | 1 | CPIC PharmGKB Reference Tables |
| CYP3A5 | 3 | Null | 0 | CPIC PharmGKB Reference Tables |
| CYP2C19 | 1 | Normal | 1 | Lesche et al., 2019 |
| CYP2C19 | 2 | Null | 0 | Lesche et al., 2019 |
| CYP2C19 | 8 | Null | 0 | CPIC PharmGKB Reference Tables |
| CYP2C19 | 15 | Normal | 1 | CPIC PharmGKB Reference Tables |
| CYP2C19 | 17 | Increased | 1.5 | Lesche et al., 2019 |
| CYP2C9 | 1 | Normal | 1 | pypgx |
| CYP2C9 | 2 | Decreased | 0.5 | pypgx |
| CYP2C9 | 3 | Null | 0 | pypgx |
| CYP2C9 | 9 | Normal | 1 | pypgx |
| CYP2C9 | 11 | Decreased | 0.5 | pypgx |
| CYP2C9 | 12 | Decreased | 0.5 | pypgx |

Supplementary Table 1. CYP pharmacogenomic star alleles called in the CardiffCOGS sample. Star allele function, and corresponding activity scores are listed, alongside sources for mapping of star alleles to activity scores.

Supplementary Table 2. Descriptive statistics for key variables.

| **Variable** | **Mean (SD)** |
|---|---|
| Chlorpromazine-Equivalent Antipsychotic Dose (mg/day) | 587.6 (390.1) |
| Age | 43.5 (11.7) |
| **Variable** | **n (%)** |
| Clozapine Use | |
| No | 370 (63%) |
| Yes | 215 (37%) |
| Anticholinergic Use | |
| No | 494 (85%) |
| Yes | 91 (16%) |
| Adherent | |
| No | 31 (5.3%) |
| Yes | 554 (94.7%) |
| Sex | |
| Male | 380 (65%) |
| Female | 205 (35%) |
| Antipsychotic Polypharmacy | |
| No | 478 (82%) |
| Yes | 107 (18%) |
| Cigarette Use | |
| No | 235 (44%) |
| Yes | 305 (56%) |
| Unknown | 45 |
| Fluoxetine Use | |
| No | 551 (94%) |
| Yes | 34 (5.8%) |
| Carbamazepine Use | |
| No | 580 (99.1%) |
| Yes | 5 (0.9%) |

Supplementary Table 2. Descriptive statistics for key variables in the analysed sample (N = 585). Mean (SD) is reported for continuous variables and n (%) is reported for categorical variables.

Supplementary Table 3 – Antipsychotic use in CardiffCOGS

| Antipsychotic | Administration | N |
|---|---|---|
| Amisulpride | Oral | 50 |
| Aripiprazole | Oral | 74 |
| Chlorpromazine | Oral | 8 |
| Clozapine | Oral | 215 |
| Flupenthixol | Oral | 7 |
| Flupenthixol | Depot | 53 |
| Fluphenazine | Oral | 1 |
| Fluphenazine | Depot | 5 |
| Haloperidol | Oral | 7 |
| Haloperidol | Depot | 8 |
| Olanzapine | Oral | 103 |
| Pimozide | Oral | 1 |
| Pipotiazine | Depot | 1 |
| Quetiapine | Oral | 52 |
| Risperidone | Oral | 40 |
| Risperidone | Depot | 41 |
| Sulpiride | Oral | 10 |
| Trifluoperazine | Oral | 2 |
| Zuclopenthixol | Oral | 2 |
| Zuclopenthixol | Depot | 11 |
| Paliperidone | Depot | 3 |

Supplementary Table 3. Frequency of currently used antipsychotics in the CardiffCOGS sample and their mode of administration.

Supplementary Table 4. CYP Pharmacogenomic star allele frequencies in CardiffCOGS

| Gene | Star Allele | Total | Frequency |
|---|---|---|---|
| *CYP1A2* | *1A | 403 | 0.344 |
| | *1C*1F | 16 | 0.014 |
| | *1F | 748 | 0.639 |
| | *1K | 3 | 0.003 |
| *CYP2D6* | *1 | 556 | 0.475 |
| | *10 | 10 | 0.009 |
| | *2 | 181 | 0.155 |
| | *35 | 50 | 0.043 |
| | *4 | 246 | 0.210 |
| | *41 | 110 | 0.094 |
| | *117 | 2 | 0.002 |
| | *17 | 5 | 0.004 |
| | *28 | 5 | 0.004 |
| | *59 | 5 | 0.004 |
| *CYP3A5* | *1 | 67 | 0.057 |
| | *3 | 1103 | 0.943 |
| *CYP2C19* | *1 | 740 | 0.632 |
| | *15 | 1 | 0.001 |
| | *17 | 255 | 0.218 |
| | *2 | 172 | 0.147 |
| | *8 | 2 | 0.002 |
| *CYP2C9* | *1 | 946 | 0.809 |
| | *2 | 135 | 0.115 |
| | *3 | 78 | 0.067 |
| | *11 | 8 | 0.007 |
| | *12 | 2 | 0.002 |

Supplementary Table 4. Total allele count, and allele frequency for CYP pharmacogenomic star alleles found in the CardiffCOGS sample.

Supplementary Table 5. Summary of CFA results

| Latent Factor | Variable | Mean | SD | Min | Max | M1 | M2 | M3 | M4 |
|---|---|---|---|---|---|---|---|---|---|
| Positive | SAPS Global Hallucinations | 1.338 | 1.852 | 0.00 | 5.00 | 0.769 | 0.874 | 0.681 | 0.858 |
| Positive | SAPS Global Delusions | 1.255 | 1.595 | 0.00 | 5.00 | 0.750 | 0.659 | 0.847 | 0.672 |
| Diminished Expressivity | SANS Global Affective Flattening | 1.383 | 1.435 | 0.00 | 5.00 | 0.872 | 0.880 | 0.859 | 0.867 |
| Diminished Expressivity | SANS Global Alogia | 1.125 | 1.475 | 0.00 | 5.00 | 0.771 | 0.764 | 0.782 | 0.775 |
| Reduced Motivation and Pleasure | SANS Global Avolition/Apathy | 1.781 | 1.327 | 0.00 | 5.00 | 0.840 | 0.839 | 0.847 | 0.846 |
| Reduced Motivation and Pleasure | SANS Global Anhedonia/Asociality | 1.853 | 1.516 | 0.00 | 5.00 | 0.760 | 0.762 | 0.754 | 0.756 |
| Disorganised | SAPS Global Positive Thought Disorder | 0.583 | 1.106 | 0.00 | 5.00 | 0.492 | *EXCL* | 0.493 | *EXCL* |
| Disorganised | SANS Inappropriate Affect | 0.080 | 0.448 | 0.00 | 4.00 | 0.526 | *EXCL* | 0.525 | *EXCL* |
| Suicide and Depression | CDSS Suicide item | 0.157 | 0.417 | 0.00 | 3.00 | 0.913 | 0.904 | *EXCL* | *EXCL* |
| Suicide and Depression | CDSS Depression item | 0.533 | 0.747 | 0.00 | 3.00 | 0.808 | 0.817 | *EXCL* | *EXCL* |
| Cognitive ability | MATRICS Problem Solving task (mazes)* | -1.513 | 1.294 | -3.99 | 1.17 | 0.653 | 0.654 | 0.652 | 0.653 |
| Cognitive ability | MATRICS Verbal Learning task (HVLT)* | -2.447 | 1.521 | -6.38 | 1.59 | 0.743 | 0.742 | 0.744 | 0.742 |
| Cognitive ability | MATRICS Visual Learning task (BVMT)* | -1.646 | 1.222 | -3.92 | 1.52 | 0.707 | 0.708 | 0.708 | 0.709 |
| Cognitive ability | MATRICS Attention (CPT)* | -1.438 | 1.228 | -5.52 | 2.25 | 0.733 | 0.732 | 0.732 | 0.730 |
| Cognitive ability | MATRICS Processing Speed* | -1.951 | 1.104 | -5.45 | 1.32 | 0.834 | 0.835 | 0.835 | 0.836 |
| Cognitive ability | MATRICS Working Memory* | -1.641 | 1.193 | -4.59 | 1.56 | 0.831 | 0.831 | 0.832 | 0.832 |
| **Goodness of Fit Measures** | | | | | | | | | |
| CFI | | | | | | 0.996 | 0.997 | 0.996 | 0.995 |
| RMSEA | | | | | | 0.020 | 0.019 | 0.024 | 0.028 |
| SRMR | | | | | | 0.053 | 0.043 | 0.050 | 0.044 |

Supplementary Table 5. Summary of full CFA results. Descriptive statistics are listed alongside factor loadings for each variable onto its latent factors across the four considered models (M1 – M4). Key fit statistics are listed below, with the best fit highlighted in red. M1 represents the maximal model with all variables included. M2 represents the maximal model without the disorganised variables. M3 represents the maximal model without the suicide/depression variables (i.e., the 5-factor model as based on Legge et al., (2021)). M4 represents the maximal model with neither the suicide/depression nor disorganised factors. EXCL = excluded; CFI = Comparative Fit Index; RMSEA = Root Mean Square Error of Approximation; SRMR = Standardised Root Mean Square Residual.  *MATRICS variables were imputed and standardised against unaffected controls.

Supplementary Table 6. Extended Results for Manuscript Table 1

| | **Positive** | | **Diminished Expressivity** | | **Reduced Motivation & Pleasure** | | **Depression & Suicide** | | **Cognition** | |
|---|---|---|---|---|---|---|---|---|---|---|
| **Predictors** | **β (SE)** | ***p*(FDR)** | **β (SE)** | ***p*(FDR)** | **β (SE)** | ***p*(FDR)** | **β (SE)** | ***p*(FDR)** | **β (SE)** | ***p*(FDR)** |
| CPZ-eq Antipsychotic Dose (mg/day) | 0.145 (0.04) | 8x10$^{-4}$ (0.004) | 0.043 (0.04) | 0.307 (0.307) | 0.056 (0.04) | 0.195 (0.244) | 0.099 (0.04) | 0.021 (0.034) | -0.12 (0.04) | 0.001 (0.004) |
| Clozapine (Yes) | -0.061 (0.09) | 0.491 (0.613) | 0.417 (0.09) | 2x10$^{-6}$ (6x10$^{-6}$) | 0.232 (0.09) | 0.009 (0.015) | -0.007 (0.09) | 0.937 (0.937) | -0.498 (0.08) | 3x10$^{-10}$ (1x10$^{-9}$) |
| Anticholinergic (Yes) | 0.081 (0.12) | 0.493 (0.493) | 0.222 (0.12) | 0.055 (0.076) | 0.272 (0.12) | 0.021 (0.053) | 0.221 (0.12) | 0.06 (0.076) | -0.345 (0.1) | 8x10$^{-4}$ (0.004) |
| Adherent (Yes) | -0.498 (0.19) | 0.008 (0.038) | -0.235 (0.18) | 0.201 (0.201) | -0.364 (0.19) | 0.051 (0.107) | -0.345 (0.19) | 0.064 (0.107) | 0.26 (0.16) | 0.11 (0.138) |
| Schizophrenia PGS | -0.041 (0.04) | 0.335 (0.558) | 0.029 (0.04) | 0.495 (0.618) | -0.006 (0.04) | 0.879 (0.879) | -0.063 (0.04) | 0.138 (0.345) | -0.1 (0.04) | 0.009 (0.043) |
| Intelligence PGS | | | | | | | | | 0.075 (0.04) | 0.091 (0.091) |
| Educational Attainment PGS | | | | | | | | | 0.13 (0.04) | 0.003 (0.003) |
| Age | -0.048 (0.04) | 0.261 (0.435) | 0.061 (0.04) | 0.147 (0.367) | 0.036 (0.04) | 0.395 (0.493) | 0.001 (0.04) | 0.985 (0.985) | -0.376 (0.04) | 2x10$^{-22}$ (1x10$^{-21}$) |
| Sex (Female) | 0.052 (0.09) | 0.547 (0.684) | -0.176 (0.09) | 0.04 (0.146) | -0.104 (0.09) | 0.232 (0.386) | 0.165 (0.09) | 0.058 (0.146) | 0.021 (0.08) | 0.783 (0.783) |
| PC1 | -0.026 (0.04) | 0.534 (0.668) | 0.009 (0.04) | 0.835 (0.835) | -0.028 (0.04) | 0.501 (0.668) | -0.065 (0.04) | 0.126 (0.631) | 0.029 (0.04) | 0.429 (0.668) |
| PC2 | -0.034 (0.04) | 0.411 (0.594) | 0.026 (0.04) | 0.52 (0.594) | 0.022 (0.04) | 0.594 (0.594) | -0.054 (0.04) | 0.192 (0.481) | -0.047 (0.04) | 0.191 (0.481) |
| PC3 | -0.062 (0.04) | 0.132 (0.658) | 0.008 (0.04) | 0.835 (0.97) | 0.008 (0.04) | 0.85 (0.97) | 0.002 (0.04) | 0.97 (0.97) | -0.018 (0.04) | 0.617 (0.97) |
| PC4 | 0.003 (0.04) | 0.942 (0.942) | -0.031 (0.04) | 0.455 (0.892) | -0.015 (0.04) | 0.713 (0.892) | -0.021 (0.04) | 0.616 (0.892) | 0.041 (0.04) | 0.264 (0.892) |
| PC5 | -0.032 (0.04) | 0.441 (0.551) | 0.064 (0.04) | 0.115 (0.315) | 0.063 (0.04) | 0.126 (0.315) | 0.034 (0.04) | 0.402 (0.551) | 0.021 (0.04) | 0.562 (0.562) |
| R2 | 0.045 | | 0.068 | | 0.042 | | 0.041 | | 0.274 | |
| Adjusted R2 | 0.025 | | 0.049 | | 0.022 | | 0.021 | | 0.256 | |
| N | 585 | | 585 | | 585 | | 585 | | 585 | |

Supplementary Table 6. Associations between medication variables with schizophrenia symptom severity and cognitive ability dimensions in CardiffCOGS, showing extended results from Table 1 in the main text. Standardised regression estimates are reported. CPZ-eq = chlorpromazine-equivalent; SE = Standard Error; PGS = Polygenic Score; PC = Genetic Principal Component.

Supplementary Table 7. Extended Results for Manuscript Table 2

| | Positive | | Diminished Expressivity | | Reduced Motivation & Pleasure | | Depression & Suicide | | Cognition | |
|---|---|---|---|---|---|---|---|---|---|---|
| **Predictors** | **β (SE)** | ***p*(FDR)** | **β (SE)** | ***p*(FDR)** | **β (SE)** | ***p*(FDR)** | **β (SE)** | ***p*(FDR)** | **β (SE)** | ***p*(FDR)** |
| CPZ-eq Antipsychotic Dose (mg/day) | 0.146 (0.04) | 7x10$^{-4}$ (0.003) | 0.035 (0.04) | 0.405 (0.405) | 0.045 (0.04) | 0.296 (0.37) | 0.098 (0.04) | 0.023 (0.039) | -0.115 (0.04) | 0.003 (0.006) |
| Clozapine (Yes) | -0.044 (0.09) | 0.623 (0.778) | 0.414 (0.09) | 3x10$^{-6}$ (8x10$^{-6}$) | 0.236 (0.09) | 0.008 (0.014) | 0.004 (0.09) | 0.961 (0.961) | -0.499 (0.08) | 4x10$^{-10}$ (2x10$^{-9}$) |
| Anticholinergic (Yes) | 0.059 (0.12) | 0.618 (0.618) | 0.209 (0.12) | 0.073 (0.104) | 0.266 (0.12) | 0.025 (0.062) | 0.205 (0.12) | 0.083 (0.104) | -0.329 (0.1) | 0.002 (0.008) |
| CYP1A2 Activity Score | -0.051 (0.04) | 0.227 (0.365) | -0.066 (0.04) | 0.116 (0.289) | -0.039 (0.04) | 0.365 (0.365) | -0.042 (0.04) | 0.321 (0.365) | 0.07 (0.04) | 0.06 (0.289) |
| CYP2D6 Activity Score | -0.022 (0.04) | 0.595 (0.744) | 0.047 (0.04) | 0.25 (0.67) | 0.046 (0.04) | 0.268 (0.67) | -0.008 (0.04) | 0.847 (0.847) | -0.02 (0.04) | 0.587 (0.744) |
| CYP3A5 Activity Score | -0.042 (0.04) | 0.319 (0.333) | -0.113 (0.04) | 0.007 (0.031) | -0.106 (0.04) | 0.012 (0.031) | -0.041 (0.04) | 0.333 (0.333) | 0.037 (0.04) | 0.321 (0.333) |
| CYP2C19 Activity Score | -0.108 (0.04) | 0.009 (0.047) | -0.003 (0.04) | 0.936 (0.936) | -0.04 (0.04) | 0.339 (0.565) | -0.079 (0.04) | 0.057 (0.143) | 0.008 (0.04) | 0.817 (0.936) |
| CYP2C9 Activity Score | -0.049 (0.04) | 0.237 (0.54) | -0.012 (0.04) | 0.764 (0.919) | 0.004 (0.04) | 0.919 (0.919) | -0.041 (0.04) | 0.324 (0.54) | -0.048 (0.04) | 0.188 (0.54) |
| Adherent (Yes) | -0.418 (0.19) | 0.026 (0.122) | -0.257 (0.19) | 0.167 (0.167) | -0.371 (0.19) | 0.051 (0.122) | -0.34 (0.19) | 0.073 (0.122) | 0.257 (0.17) | 0.121 (0.152) |
| Schizophrenia PGS | -0.044 (0.04) | 0.3 (0.501) | 0.021 (0.04) | 0.615 (0.73) | -0.015 (0.04) | 0.73 (0.73) | -0.063 (0.04) | 0.144 (0.36) | -0.092 (0.04) | 0.017 (0.083) |
| Intelligence PGS | | | | | | | | | 0.084 (0.04) | 0.06 (0.06) |
| Educational Attainment PGS | | | | | | | | | 0.123 (0.04) | 0.006 (0.006) |
| Age | -0.052 (0.04) | 0.218 (0.363) | 0.059 (0.04) | 0.162 (0.363) | 0.033 (0.04) | 0.444 (0.555) | -0.005 (0.04) | 0.91 (0.91) | -0.375 (0.04) | 9x10$^{-22}$ (4x10$^{-21}$) |
| Sex (Female) | 0.053 (0.09) | 0.541 (0.676) | -0.197 (0.09) | 0.023 (0.115) | -0.126 (0.09) | 0.151 (0.252) | 0.147 (0.09) | 0.094 (0.236) | 0.03 (0.08) | 0.698 (0.698) |
| PC1 | -0.019 (0.04) | 0.651 (0.724) | 0.02 (0.04) | 0.628 (0.724) | -0.015 (0.04) | 0.724 (0.724) | -0.057 (0.04) | 0.18 (0.724) | 0.029 (0.04) | 0.433 (0.724) |
| PC2 | -0.032 (0.04) | 0.442 (0.665) | 0.022 (0.04) | 0.595 (0.665) | 0.018 (0.04) | 0.665 (0.665) | -0.052 (0.04) | 0.209 (0.539) | -0.045 (0.04) | 0.216 (0.539) |
| PC3 | -0.048 (0.04) | 0.248 (0.728) | 0.022 (0.04) | 0.589 (0.728) | 0.023 (0.04) | 0.578 (0.728) | 0.014 (0.04) | 0.728 (0.728) | -0.018 (0.04) | 0.615 (0.728) |
| PC4 | 0.007 (0.04) | 0.866 (0.961) | -0.016 (0.04) | 0.703 (0.961) | -0.002 (0.04) | 0.961 (0.961) | -0.018 (0.04) | 0.676 (0.961) | 0.036 (0.04) | 0.33 (0.961) |
| PC5 | -0.03 (0.04) | 0.466 (0.582) | 0.076 (0.04) | 0.064 (0.24) | 0.069 (0.04) | 0.096 (0.24) | 0.036 (0.04) | 0.388 (0.582) | 0.012 (0.04) | 0.751 (0.751) |
| R2 | 0.060 | | 0.084 | | 0.056 | | 0.051 | | 0.280 | |
| Adjusted R2 | 0.032 | | 0.056 | | 0.027 | | 0.022 | | 0.256 | |
| N | 578 | | 578 | | 578 | | 578 | | 578 | |

Supplementary Table 7. Associations between medication and pharmacogenomic variables with schizophrenia symptom severity and cognitive ability dimensions in CardiffCOGS, showing extended results from Table 2 in the main text. Standardised regression estimates are reported. CPZ-eq = chlorpromazine-equivalent; SE = Standard Error; PGS = Polygenic Score; PC = Genetic Principal Component.

Supplementary Table 8. Extended Results for Manuscript Table 3

| | Positive (Clozapine Subgroup) | | Diminished Expressivity (Clozapine Subgroup) | | Reduced Motivation & Pleasure (Clozapine Subgroup) | | Depression & Suicide (Clozapine Subgroup) | | Cognition (Clozapine Subgroup) | |
|---|---|---|---|---|---|---|---|---|---|---|
| **Predictors** | **β (SE)** | ***p*(FDR)** | **β (SE)** | ***p*(FDR)** | **β (SE)** | ***p*(FDR)** | **β (SE)** | ***p*(FDR)** | **β (SE)** | ***p*(FDR)** |
| CPZ-eq Antipsychotic Dose (mg/day) | 0.02 (0.07) | 0.786 (0.786) | 0.077 (0.07) | 0.273 (0.683) | 0.03 (0.07) | 0.673 (0.786) | 0.05 (0.07) | 0.471 (0.785) | -0.147 (0.06) | 0.02 (0.1) |
| Anticholinergic (Yes) | -0.076 (0.2) | 0.704 (0.88) | 0.023 (0.19) | 0.907 (0.907) | 0.137 (0.2) | 0.486 (0.81) | 0.189 (0.19) | 0.327 (0.81) | -0.274 (0.17) | 0.109 (0.547) |
| CYP1A2 Activity Score | -0.083 (0.07) | 0.242 (0.404) | -0.108 (0.07) | 0.117 (0.293) | -0.041 (0.07) | 0.556 (0.556) | -0.059 (0.07) | 0.387 (0.484) | 0.17 (0.06) | 0.005 (0.026) |
| CYP2D6 Activity Score | -0.028 (0.07) | 0.695 (0.836) | 0.071 (0.07) | 0.315 (0.836) | 0.064 (0.07) | 0.369 (0.836) | -0.039 (0.07) | 0.574 (0.836) | -0.013 (0.06) | 0.836 (0.836) |
| CYP3A5 Activity Score | -0.082 (0.07) | 0.251 (0.251) | -0.12 (0.07) | 0.085 (0.198) | -0.103 (0.07) | 0.147 (0.198) | -0.126 (0.07) | 0.07 (0.198) | 0.087 (0.06) | 0.158 (0.198) |
| CYP2C19 Activity Score | -0.05 (0.07) | 0.481 (0.843) | 0.014 (0.07) | 0.843 (0.843) | 0.014 (0.07) | 0.84 (0.843) | 0.052 (0.07) | 0.444 (0.843) | 0.023 (0.06) | 0.699 (0.843) |
| CYP2C9 Activity Score | -0.041 (0.07) | 0.564 (0.62) | -0.164 (0.07) | 0.019 (0.095) | -0.121 (0.07) | 0.086 (0.215) | -0.098 (0.07) | 0.155 (0.258) | 0.03 (0.06) | 0.62 (0.62) |
| Adherent (Yes) | -0.168 (0.52) | 0.749 (0.749) | -0.306 (0.51) | 0.549 (0.749) | -0.411 (0.52) | 0.429 (0.749) | -0.36 (0.51) | 0.479 (0.749) | 0.226 (0.45) | 0.615 (0.749) |
| Schizophrenia PGS | -0.021 (0.07) | 0.772 (0.772) | 0.022 (0.07) | 0.748 (0.772) | -0.034 (0.07) | 0.632 (0.772) | -0.135 (0.07) | 0.053 (0.229) | -0.108 (0.06) | 0.092 (0.229) |
| Intelligence PGS | | | | | | | | | 0.14 (0.08) | 0.076 (0.076) |
| Educational Attainment PGS | | | | | | | | | 0.083 (0.08) | 0.279 (0.279) |
| Age | -0.135 (0.07) | 0.06 (0.15) | 0.057 (0.07) | 0.41 (0.683) | 0.017 (0.07) | 0.813 (0.992) | 0.001 (0.07) | 0.992 (0.992) | -0.398 (0.06) | $6x10^{-10}$ ($3x10^{-9}$) |
| Sex (Female) | 0.236 (0.15) | 0.112 (0.279) | -0.192 (0.14) | 0.182 (0.304) | -0.067 (0.15) | 0.649 (0.812) | 0.245 (0.14) | 0.089 (0.279) | 0.008 (0.13) | 0.948 (0.948) |
| PC1 | -0.036 (0.08) | 0.631 (0.631) | -0.11 (0.07) | 0.131 (0.164) | -0.114 (0.07) | 0.127 (0.164) | -0.165 (0.07) | 0.024 (0.121) | 0.102 (0.06) | 0.111 (0.164) |
| PC2 | -0.087 (0.15) | 0.566 (0.566) | -0.197 (0.15) | 0.181 (0.302) | -0.232 (0.15) | 0.123 (0.302) | -0.316 (0.15) | 0.032 (0.16) | 0.132 (0.13) | 0.305 (0.381) |
| PC3 | 0.018 (0.29) | 0.951 (0.951) | 0.377 (0.28) | 0.181 (0.302) | 0.331 (0.29) | 0.247 (0.309) | 0.46 (0.28) | 0.102 (0.254) | -0.41 (0.25) | 0.097 (0.254) |
| PC4 | -0.024 (0.28) | 0.932 (0.932) | -0.448 (0.27) | 0.1 (0.126) | -0.469 (0.28) | 0.091 (0.126) | -0.467 (0.27) | 0.086 (0.126) | 0.396 (0.24) | 0.097 (0.126) |
| PC5 | 0.053 (0.1) | 0.61 (0.763) | -0.136 (0.1) | 0.178 (0.444) | -0.216 (0.1) | 0.036 (0.182) | 0.052 (0.1) | 0.601 (0.763) | 0.019 (0.09) | 0.833 (0.833) |
| R2 | 0.063 | | 0.114 | | 0.082 | | 0.122 | | 0.335 | |
| Adjusted R2 | -0.013 | | 0.043 | | 0.008 | | 0.051 | | 0.273 | |
| N | 215 | | 215 | | 215 | | 215 | | 215 | |

Supplementary Table 8. Associations between medication and pharmacogenomic variables with schizophrenia symptom severity and cognitive ability dimensions in CardiffCOGS participants currently prescribed clozapine, showing extended results from Table 3 in the main text. Standardised regression estimates are reported. CPZ-eq = chlorpromazine-equivalent; SE = Standard Error; PGS = Polygenic Score; PC = Genetic Principal Component.

Supplementary Table 9. Unadjusted estimates for pharmacogenomic variables in total sample.

| | Positive | | Diminished Expressivity | | Reduced Motivation & Pleasure | | Depression & Suicide | | Cognition | |
|---|---|---|---|---|---|---|---|---|---|---|
| **Enzyme** | **r** | ***p*** | **r** | ***p*** | **r** | ***p*** | **r** | ***p*** | **r** | ***P*** |
| CYP1A2 Activity Score | -0.052 | 0.209 | -0.050 | 0.231 | -0.026 | 0.525 | -0.044 | 0.290 | 0.092 | 0.026 |
| CYP2D6 Activity Score | 0.004 | 0.919 | 0.048 | 0.245 | 0.048 | 0.248 | 0.014 | 0.744 | -0.063 | 0.128 |
| CYP3A5 Activity Score | -0.046 | 0.265 | -0.098 | 0.018 | -0.095 | 0.022 | -0.040 | 0.333 | 0.025 | 0.548 |
| CYP2C19 Activity Score | -0.114 | 0.006 | -0.003 | 0.937 | -0.045 | 0.281 | -0.085 | 0.041 | 0.017 | 0.687 |
| CYP2C9 Activity Score | -0.050 | 0.233 | -0.029 | 0.486 | -0.009 | 0.821 | -0.046 | 0.268 | -0.026 | 0.533 |

Supplementary Table 9. Unadjusted estimates for pharmacogenomic variables against schizophrenia phenotype dimensions. Table shows Pearson's correlation coefficient and *p* value for each dimension-enzyme pairing.

Supplementary Table 10. Unadjusted estimates for pharmacogenomic variables in the subgroup of participants prescribed clozapine.

| | Positive | | Diminished Expressivity | | Reduced Motivation & Pleasure | | Depression & Suicide | | Cognition | |
|---|---|---|---|---|---|---|---|---|---|---|
| **Enzyme** | **r** | ***p*** | **r** | ***p*** | **r** | ***p*** | **r** | ***p*** | **r** | ***P*** |
| CYP1A2 Activity Score | -0.055 | 0.422 | -0.102 | 0.137 | -0.035 | 0.613 | -0.040 | 0.561 | 0.205 | 0.002 |
| CYP2D6 Activity Score | -0.014 | 0.844 | 0.114 | 0.094 | 0.102 | 0.135 | 0.025 | 0.711 | -0.110 | 0.107 |
| CYP3A5 Activity Score | -0.084 | 0.221 | -0.113 | 0.100 | -0.105 | 0.125 | -0.123 | 0.072 | 0.068 | 0.319 |
| CYP2C19 Activity Score | -0.066 | 0.337 | 0.017 | 0.806 | 0.009 | 0.890 | 0.038 | 0.575 | -0.026 | 0.705 |
| CYP2C9 Activity Score | -0.044 | 0.517 | -0.186 | 0.006 | -0.142 | 0.037 | -0.138 | 0.043 | 0.089 | 0.195 |

Supplementary Table 10. Unadjusted estimates for pharmacogenomic variables against Schizophrenia Phenotype Dimensions. Table shows Pearson's correlation coefficient and *p* value for each dimension-enzyme pairing dimensions in CardiffCOGS participants currently prescribed clozapine.

Supplementary Table 11. Sensitivity analyses accounting for potential fluoxetine-induced CYP2C19 phenoconversion.

| | **Positive CYP2C19 – Fluoxetine Interaction** | | **Positive CYP2C19 Phenoconversion-Corrected Activity Scores** | |
|---|---|---|---|---|
| **Predictors** | **β (SE)** | ***p*** | **β (SE)** | ***p*** |
| CPZ-eq Antipsychotic Dose (mg/day) | 0.135 (0.04) | 0.002 | 0.145 (0.04) | $7x10^{-4}$ |
| Clozapine (Yes) | -0.056 (0.09) | 0.522 | -0.058 (0.09) | 0.515 |
| Anticholinergic (Yes) | 0.081 (0.12) | 0.487 | 0.077 (0.12) | 0.513 |
| CYP2C19 Activity Score | -0.09 (0.04) | 0.037 | | |
| Fluoxetine (Yes) | -0.363 (0.18) | 0.04 | | |
| CYP2C19-Fluoxetine interaction | -0.123 (0.15) | 0.408 | | |
| Phenoconversion-Corrected CYP2C19 Activity Score | | | -0.022 (0.04) | 0.6 |
| Adherent (Yes) | -0.517 (0.19) | 0.005 | -0.491 (0.19) | 0.009 |
| Schizophrenia PGS | -0.045 (0.04) | 0.289 | -0.042 (0.04) | 0.321 |
| Age | -0.056 (0.04) | 0.186 | -0.048 (0.04) | 0.262 |
| Sex (Female) | 0.07 (0.09) | 0.419 | 0.05 (0.09) | 0.566 |
| PC1 | -0.027 (0.04) | 0.525 | -0.025 (0.04) | 0.549 |
| PC2 | -0.019 (0.04) | 0.643 | -0.036 (0.04) | 0.392 |
| PC3 | -0.053 (0.04) | 0.196 | -0.061 (0.04) | 0.139 |
| PC4 | 0.003 (0.04) | 0.95 | 0.003 (0.04) | 0.949 |
| PC5 | -0.034 (0.04) | 0.401 | -0.033 (0.04) | 0.428 |
| R2 | 0.062 | | 0.045 | |
| Adjusted R2 | 0.037 | | 0.024 | |
| N | 585 | | 585 | |

Supplementary Table 11. Sensitivity analyses to account for the potential of fluoxetine to inhibit CYP2C19 activity. This is accounted for through a control (fluoxetine yes/no) covariate, or by inclusion of a phenoconversion activity score (activity score * 0 if participant reports fluoxetine). Sensitivity analyses were performed only in the positive dimension where an association with CYP2C19 was previously observed.

Supplementary Table 12. Sensitivity analyses accounting for potential carbamazepine-induced CYP3A5 phenoconversion.

| | Diminished Expressivity CYP3A5 – Carbamazepine Covariate | | Reduced Motivation & Pleasure CYP3A5 – Carbamazepine Interaction | |
|---|---|---|---|---|
| **Predictors** | **β (SE)** | ***p*** | **β (SE)** | ***p*** |
| CPZ-eq Antipsychotic Dose (mg/day) | 0.043 (0.04) | 0.303 | 0.056 (0.04) | 0.192 |
| Clozapine (Yes) | 0.428 (0.09) | $1x10^{-6}$ | 0.242 (0.09) | 0.006 |
| Anticholinergic (Yes) | 0.236 (0.12) | 0.041 | 0.284 (0.12) | 0.015 |
| CYP3A5 Activity Score | -0.106 (0.04) | 0.01 | -0.1 (0.04) | 0.018 |
| Carbamazepine (Yes) | 0.644 (0.44) | 0.142 | 0.597 (0.44) | 0.18 |
| Adherent (Yes) | -0.229 (0.18) | 0.21 | -0.359 (0.19) | 0.053 |
| Schizophrenia PGS | 0.029 (0.04) | 0.492 | -0.006 (0.04) | 0.88 |
| Age | 0.065 (0.04) | 0.12 | 0.04 (0.04) | 0.345 |
| Sex (Female) | -0.181 (0.09) | 0.034 | -0.109 (0.09) | 0.21 |
| PC1 | 0.024 (0.04) | 0.573 | -0.015 (0.04) | 0.732 |
| PC2 | 0.02 (0.04) | 0.631 | 0.016 (0.04) | 0.703 |
| PC3 | 0.02 (0.04) | 0.629 | 0.018 (0.04) | 0.659 |
| PC4 | -0.018 (0.04) | 0.663 | -0.003 (0.04) | 0.937 |
| PC5 | 0.064 (0.04) | 0.112 | 0.063 (0.04) | 0.124 |
| R2 | 0.083 | | 0.054 | |
| Adjusted R2 | 0.060 | | 0.031 | |
| N | 585 | | 585 | |

Supplementary Table 12. Sensitivity analysis to account for the potential of carbamazepine to induce CYP3A5 activity. This is accounted for through a control (carbamazepine yes/no) covariate for both symptom dimensions (i.e., diminished expressivity, reduced motivation and pleasure) where an association with CYP3A5 activity score was previously observed.

Supplementary Table 13. Sensitivity analyses accounting for potential smoking-induced CYP1A2 phenoconversion.

| | Cognitive (Clozapine Subgroup) CYP1A2 – Cigarette Interaction | | Cognitive (Clozapine Subgroup) CYP1A2 Phenoconversion-Corrected Activity Scores | |
|---|---|---|---|---|
| **Predictors** | **β (SE)** | ***p*** | **β (SE)** | ***p*** |
| CPZ-eq Antipsychotic Dose (mg/day) | -0.133 (0.06) | 0.038 | -0.147 (0.06) | 0.024 |
| Anticholinergic (Yes) | -0.352 (0.17) | 0.043 | -0.375 (0.18) | 0.034 |
| CYP1A2 Activity Score | 0.19 (0.1) | 0.06 | | |
| Cigarettes (Yes) | -0.29 (0.13) | 0.026 | | |
| CYP1A2-Smoking interaction | -0.066 (0.13) | 0.618 | | |
| Phenoconversion-Corrected CYP1A2 Activity Score | | | -0.022 (0.06) | 0.739 |
| Adherent (Yes) | 0.206 (0.44) | 0.644 | 0.207 (0.45) | 0.648 |
| Schizophrenia PGS | -0.092 (0.07) | 0.165 | -0.102 (0.07) | 0.133 |
| Intelligence PGS | 0.147 (0.08) | 0.078 | 0.14 (0.08) | 0.094 |
| Educational Attainment PGS | 0.032 (0.08) | 0.704 | 0.074 (0.08) | 0.367 |
| Age | -0.401 (0.06) | $8x10^{-10}$ | -0.405 (0.06) | $9x10^{-10}$ |
| Sex (Female) | 0.029 (0.13) | 0.823 | 0.066 (0.13) | 0.617 |
| PC1 | 0.094 (0.06) | 0.143 | 0.09 (0.06) | 0.165 |
| PC2 | 0.138 (0.13) | 0.293 | 0.178 (0.13) | 0.18 |
| PC3 | -0.429 (0.25) | 0.091 | -0.488 (0.26) | 0.058 |
| PC4 | 0.403 (0.25) | 0.102 | 0.482 (0.25) | 0.054 |
| PC5 | -0.018 (0.09) | 0.848 | 0.008 (0.09) | 0.931 |
| R2 | 0.339 | | 0.301 | |
| Adjusted R2 | 0.284 | | 0.251 | |
| N | 197 | | 197 | |

Supplementary Table 13. Sensitivity analysis to account for the potential of smoking to influence CYP1A2 activity. This is accounted for through a control (smoking yes/no) covariate, or by inclusion of a phenoconversion activity score (activity score * 1.5 if participant is a smoker). Sensitivity analyses were performed only in the cognitive dimension where an association with CYP1A2 activity score was previously observed.

Supplementary Table 14. Alternative Analyses using NART IQ as a proxy of premorbid intelligence

| | Medication Model | | Medication & Pharmacogenomic Model | | Medication & Pharmacogenomic Model (Clozapine Subgroup) | |
|---|---|---|---|---|---|---|
| **Predictors** | **β (SE)** | ***p*** | **β (SE)** | ***p*** | **β (SE)** | ***p*** |
| CPZ-eq Antipsychotic Dose (mg/day) | -0.071 (0.03) | 0.03 | -0.066 (0.03) | 0.047 | -0.134 (0.05) | 0.012 |
| Clozapine current (Yes) | -0.339 (0.07) | $7x10^{-7}$ | -0.339 (0.07) | $1x10^{-6}$ | | |
| Anticholinergic (Yes) | -0.276 (0.09) | 0.002 | -0.271 (0.09) | 0.003 | -0.187 (0.15) | 0.203 |
| CYP1A2 Activity Score | | | 0.025 (0.03) | 0.433 | 0.066 (0.05) | 0.221 |
| CYP2D6 Activity Score | | | -0.037 (0.03) | 0.25 | -0.02 (0.05) | 0.7 |
| CYP3A5 Activity Score | | | 0.039 (0.03) | 0.221 | 0.082 (0.05) | 0.117 |
| CYP2C19 Activity Score | | | 0.018 (0.03) | 0.57 | 0.005 (0.05) | 0.917 |
| CYP2C9 Activity Score | | | -0.025 (0.03) | 0.435 | 0.031 (0.05) | 0.553 |
| NART IQ | 0.475 (0.03) | $3x10^{-42}$ | 0.474 (0.03) | $7x10^{-41}$ | 0.454 (0.05) | $5x10^{-15}$ |
| Adherent (Yes) | 0.254 (0.14) | 0.073 | 0.26 (0.14) | 0.073 | 0.056 (0.38) | 0.881 |
| Schizophrenia PGS | -0.063 (0.03) | 0.049 | -0.059 (0.03) | 0.068 | -0.097 (0.05) | 0.063 |
| Age | -0.442 (0.03) | $3x10^{-42}$ | 0.438 (0.03) | $8x10^{-36}$ | -0.481 (0.05) | $1x10^{-16}$ |
| Sex (Female) | 0.103 (0.07) | 0.118 | 0.112 (0.07) | 0.094 | 0.053 (0.11) | 0.63 |
| PC1 | 0.074 (0.03) | 0.023 | 0.069 (0.03) | 0.038 | 0.059 (0.05) | 0.276 |
| PC2 | -0.023 (0.03) | 0.446 | -0.022 (0.03) | 0.474 | 0.161 (0.11) | 0.157 |
| PC3 | -0.035 (0.03) | 0.254 | -0.04 (0.03) | 0.199 | -0.509 (0.22) | 0.021 |
| PC4 | 0.027 (0.03) | 0.371 | 0.022 (0.03) | 0.483 | 0.398 (0.21) | 0.061 |
| PC5 | 0.009 (0.03) | 0.778 | 0.004 (0.03) | 0.904 | -0.088 (0.08) | 0.257 |
| R2 | 0.469 | | 0.470 | | 0.510 | |
| Adjusted R2 | 0.457 | | 0.452 | | 0.465 | |
| N | 558 | | 551 | | 202 | |

Supplementary Table 14. Associations between medication and pharmacogenomic variables with the cognitive ability dimensions in CardiffCOGS. NART IQ is included as a proxy of premorbid intelligence in place of the Intelligence PGS and Educational Attainment PGS used in the analyses presented in the main text. Standardised regression estimates are reported. CPZ-eq = chlorpromazine-equivalent; SE = Standard Error; PGS = Polygenic Score; PC = Genetic Principal Component.

STrengthening the REporting of Genetic Association studies (STREGA) reporting recommendations, extended from STROBE Statement

| Item | Item no | STROBE Guideline | Extension for Genetic Association Studies (STREGA) | Page no |
|---|---|---|---|---|
| **Title and Abstract** | 1 | (a) Indicate the study's design with a commonly used term in the title or the abstract. | | Manuscript – title/abstract (page 1 – 2) |
| | | (b) Provide in the abstract an informative and balanced summary of what was done and what was found. | | Manuscript – Abstract (page 2) |
| **Introduction** | | | | |
| *Background rationale* | 2 | Explain the scientific background and rationale for the investigation being reported. | | Manuscript – introduction (page 3) |
| *Objectives* | 3 | State specific objectives, including any pre-specified hypotheses | ***State if the study is the first report of a genetic association, a replication effort, or both.*** | Manuscript – introduction (page 3) |
| **Methods** | | | | |
| *Study design* | 4 | Present key elements of study design early in the paper. | | Manuscript – introduction and methods (page 3 – 6) |
| *Setting* | 5 | Describe the setting, locations and relevant dates, including periods of recruitment, exposure, follow-up and data collection. | | Manuscript – methods (page 4) |
| *Participants* | 6 | (a) **Cohort study –** Give the eligibility criteria, and the sources and methods of selection of participants. Describe methods of follow-up.<br><br>**Case–control study –** Give the eligibility criteria, and the sources and methods of case ascertainment and control selection. Give the rationale for the choice of cases and controls.<br><br>**Cross-sectional study –** Give the eligibility criteria, and the sources and methods of selection of participants. | ***Give information on the criteria and methods for selection of subsets of participants from a larger study, when relevant.*** | Manuscript – methods (page 4) |
| | | **(b) Cohort study** – For matched studies, give matching criteria and number of exposed and unexposed.<br><br>**Case–control study –** For matched studies, give matching criteria and the number of controls per case. | | |
| *Variables* | 7 | (a) Clearly define all outcomes, exposures, predictors, potential confounders, and effect modifiers. Give diagnostic criteria, if applicable. | ***(b) Clearly define genetic exposures (genetic variants) using a widely –used nomenclature system. Identify variables likely to be associated with population stratification (confounding by ethnic origin).*** | Manuscript – methods (page 4– 5) Supplementary materials (page 3) |
| *Data sources measurement* | 8* | (a) For each variable of interest, give sources of data and details of methods of assessment (measurement). | ***(b) Describe laboratory methods, including source and storage of DNA, genotyping methods and platforms (including the allele calling algorithm used, and its version), error rates and call rates.*** | Manuscript – methods (page 3 – 4) |

| | | | | |
|---|---|---|---|---|
| | | Describe comparability of assessment methods if there is more than one group. | ***State the laboratory /centre where genotyping was done. Describe comparability of laboratory methods if there is more than one group. Specify whether genotypes were assigned using all of the data from the study simultaneously or in smaller batches.*** | Supplementary materials (page 3) |
| *Bias* | 9 | (a) Describe any efforts to address potential sources of bias. | ***(b) For quantitative outcome variables, specify if any investigation of potential bias resulting from pharmacotherapy was undertaken. If relevant, describe the nature and magnitude of the potential bias, and explain what approach was used to deal with this.*** | Manuscript – methods (page 4) Supplementary materials (page 3 – 6) |
| *Study size* | 10 | Explain how the study size was arrived at. | | Manuscript – methods (page 4 – 6) Supplementary materials (Supplementary Figure 1) |
| *Quantitative variables* | 11 | Explain how quantitative variables were handled in the analyses. If applicable, describe which groupings were chosen, and why. | ***If applicable, describe how effects of treatment were dealt with.*** | Manuscript – methods (page 4 - 6) |
| *Statistical methods* | 12 | (a) Describe all statistical methods, including those used to control for confounding. | ***State software version used and options (or settings) chosen.*** | Manuscript – methods (page 5) |
| | | (b) Describe any methods used to examine subgroups and interactions. | | Manuscript – methods (page 5 – 6) Supplementary materials (page 3 – 6) |
| | | (c) Explain how missing data were addressed. | | |
| | | (d) **Cohort study –** If applicable, explain how loss to follow-up was addressed.<br><br>**Case–control study –** If applicable, explain how matching of cases and controls was addressed.<br><br>**Cross-sectional study –** If applicable, describe analytical methods taking account of sampling strategy. | | |
| | | (e) Describe any sensitivity analyses. | | |
| | | | ***(f) State whether Hardy- Weinberg equilibrium was considered and, if so, how.*** | Supplementary materials (page 3) |
| | | | ***(g) Describe any methods used for inferring genotypes or haplotypes.*** | Manuscript – methods (page 3 – 4) Supplementary materials (page 3) |
| | | | ***(h) Describe any methods used to assess or address population stratification.*** | Manuscript – methods (page 5) |
| | | | ***(i) Describe any methods used to address multiple comparisons or to control risk of false positive findings.*** | Manuscript – results (page 6) |
| | | | ***(j) Describe any methods used to address and correct for relatedness among subjects.*** | Supplementary materials (page 3) |
| **Results** | | | | |
| *Participants* | 13* | (a) Report the numbers of individuals at each stage of the study – e.g. numbers potentially eligible, examined for eligibility, confirmed eligible, included in the study, completing follow-up and analysed. | ***Report numbers of individuals in whom genotyping was attempted and numbers of individuals in whom genotyping was successful.*** | Supplementary materials (Supplementary Figure 1) |

| | | | | |
|---|---|---|---|---|
| | | (b) Give reasons for non-participation at each stage. | | |
| | | (c) Consider use of a flow diagram. | | |
| *Descriptive data* | 14* | (a) Give characteristics of study participants (e.g. demographic, clinical, social) and information on exposures and potential confounders. | ***Consider giving information by genotype.*** | Supplementary materials (Supplementary Tables 2 - 3) |
| | | (b) Indicate the number of participants with missing data for each variable of interest. | | |
| | | (c) **Cohort study –** Summarize follow-up time, e.g. average and total amount. | | |
| *Outcome data* | 15* | **Cohort study –** Report numbers of outcome events or summary measures over time. | ***Report outcomes (phenotypes) for each genotype category over time*** | Manuscript – results (page 6 – 7) Supplementary materials (Supplementary Table 5) |
| | | **Case–control study –** Report numbers in each exposure category, or summary measures of exposure. | ***Report numbers in each genotype category*** | |
| | | **Cross-sectional study –** Report numbers of outcome events or summary measures. | ***Report outcomes (phenotypes) for each genotype category*** | |
| *Main results* | 16 | (a) Give unadjusted estimates and, if applicable, confounder-adjusted estimates and their precision (e.g. 95% confidence intervals). Make clear which confounders were adjusted for and why they were included. | | Manuscript – results (page 6 – 7) and Tables 1 – 3. Supplementary materials (Supplementary Tables 6 – 14) |
| | | (b) Report category boundaries when continuous variables were categorized. | | |
| | | (c) If relevant, consider translating estimates of relative risk into absolute risk for a meaningful time period. | | |
| | | | ***(d) Report results of any adjustments for multiple comparisons.*** | Manuscript – results (page 6 – 7) and Tables 1 – 3. |
| *Other analyses* | 17 | (a) Report other analyses done – e.g. analyses of subgroups and interactions, and sensitivity analyses. | | Manuscript – results (page 6 – 7) Supplementary materials (page 5 – 6) |
| | | | ***(b) If numerous genetic exposures (genetic variants) were examined, summarize results from all analyses undertaken.*** | Manuscript – Table 2 – 3 Supplementary materials (Supplementary Table 7 – 8) |
| | | | ***(c) If detailed results are available elsewhere, state how they can be accessed.*** | Supplementary materials (Supplementary Tables 6 – 13) |
| **Discussion** | | | | |
| *Key results* | 18 | Summarize key results with reference to study objectives. | | Manuscript – discussion (page 7) |
| *Limitations* | 19 | Discuss limitations of the study, taking into account sources of potential bias or imprecision. Discuss both direction and magnitude of any potential bias. | | Manuscript – discussion (page 7 – 810) |
| *Interpretation* | 20 | Give a cautious overall interpretation of results considering objectives, limitations, multiplicity of | | Manuscript – discussion (page 7 – 10) |

| | | | |
|---|---|---|---|
| | | analyses, results from similar studies, and other relevant evidence. | |
| *Generalizability* | 21 | Discuss the generalizability (external validity) of the study results. | Manuscript – discussion (page 7 – 10) |
| **Other information** | | | |
| *Funding* | 22 | Give the source of funding and the role of the funders for the present study and, if applicable, for the original study on which the present article is based. | Manuscript – abstract (page 2) |
